# Longitudinal trajectories of ADHD symptoms: relationships to early-childhood predictors and later outcomes

**DOI:** 10.64898/2026.10.05.26364348

**Authors:** Emily Rounds, Michael Kozlowski, Shane McCormack, Peter Ryabinin, Elizabeth Nousen, Jessica Tipsord, Nathan F. Dieckmann, Sarah L. Karalunas, Megan M. Herting, Stephen V. Faraone, Molly Nikolas, Joel T. Nigg, Michael A. Mooney

**Author notes:** Corresponding Author: Michael A. Mooney, Ph.D. Department of Psychiatry, Oregon Health & Science University, Portland, Oregon, USA.

## Abstract

**Background:** ADHD symptoms follow heterogeneous trajectories across childhood and adolescence, with important implications for later functioning. Early-life factors, including perinatal health, family environment, cognition, socioeconomic conditions, and community exposures, may contribute to this variability. The present study examined (1) early-life predictors of ADHD symptom trajectories, and (2) ADHD symptom trajectories as predictors of later substance use, suicidality, and new-onset psychiatric disorders.

**Methods:** Growth models were used to fit longitudinal trajectories of inattention, hyperactivity, and irritability measures in two independent samples: the Oregon ADHD-1000 cohort (N=1,215; 49.17% ADHD; 39.9% female, ages 7-23) and a subsample of the Adolescent Brain Cognitive Development (ABCD) study (N=1,252; 33.3% ADHD; 31.6% female, ages 9-16). Regression analyses examined multiple risk factors as predictors of symptom growth parameters as well as associations between symptom trajectories and later adverse outcomes.

**Results:** In both cohorts, growth models fit well (CFI > .96, RMSEA < .04), with all symptoms decreasing over time (*p*s < .001) and hyperactivity declining significantly faster than inattention. ADHD polygenic risk, family conflict, parenting, and cognition were associated with symptom intercepts in both cohorts (*p*s < .001). In Oregon ADHD-1000, poorer baseline working memory and processing speed predicted slower declines in inattention (*p*s < .001), but these effects did not replicate in ABCD; no other early-life factors predicted symptom trajectories. Increasing irritability trajectories predicted new disorder onset in both cohorts (ORs = 1.73–2.09, ps < .001), while more positive inattention slopes predicted new disorder onset in ABCD (OR = 3.07, p < .001) and showed a weaker association in Oregon ADHD-1000 (OR = 1.50, p = .007).

**Conclusion:** While several early risk factors were associated with ADHD symptoms initially, none robustly predicted symptom trajectories over time. However, increasing irritability and inattention were robustly associated with increased risk for new disorder onsets later in adolescence.

**Key points:**

- ADHD symptoms decrease over time on average, with hyperactivity demonstrating the highest rate of decline.
- ADHD polygenic risk, family conflict, parental stress, parenting style, and cognition were strongly associated with symptom levels at intercept, but no replicable associations between early-life risk factors and change in symptoms over time (slopes) were observed.
- Irritability slope was robustly associated with new disorder onsets.

## INTRODUCTION

Attention-deficit/hyperactivity disorder (ADHD) is characterized by substantial heterogeneity in both clinical presentation and developmental course (Faraone et al., 2024; Luo, Weibman, Halperin, & Li, 2019). Although hyperactivity/impulsivity typically declines with age, inattention and associated impairment often persist into adulthood (Biederman, Mick, & Faraone, 2000; Cortese et al., 2025; Sasser, Kalvin, & Bierman, 2016). Importantly, these average developmental patterns obscure considerable individual variability in symptom change. Further, it remains unclear what factors *prospectively* predict individual-level patterns of symptom change and how these varying patterns are related to differential outcomes. Identifying factors that predict this variability—and whether different symptom trajectories confer differential risk for later adverse outcomes—is critical for understanding the clinical course of ADHD.

### Heterogeneity in ADHD symptom change

Longitudinal studies consistently demonstrate substantial heterogeneity in ADHD symptom trajectories. Studies examining diagnostic status change or latent grouping approaches generally identify youth with persistently elevated, declining, and consistently low symptoms, although the number and characteristics of trajectory groups vary considerably across studies (Agnew-Blais et al., 2016, 2020, 2021; Karalunas et al., 2017; Y.-S. Lee, Sprong, Shrestha, Smeltzer, & Hollender, 2025; Sasser et al., 2016; Willoughby, Williams, Mills-Koonce, & Blair, 2020). Although these approaches characterize common patterns of symptom change, considerable variability remains among individuals within trajectory groups. Consistent with this, longitudinal modeling across multiple cohorts has demonstrated overall symptom declines alongside substantial individual differences in rates of change (Wootton et al., 2022). Modeling individual-level trajectories may therefore be most informative in understanding both prospective predictors of clinical course and diverse outcomes associated with ADHD.

### Prospective predictors of trajectory

ADHD trajectories may be influenced by multiple early-life factors, including genetic liability, early environmental exposures, cognitive functioning, and psychosocial context (Agnew-Blais et al., 2021; Brinksma et al., 2023; Mooney et al., 2023; Nigg et al., 2018; Thapar & Rice, 2021). Genetic risk and adverse family or environmental factors have been associated with persistent ADHD, although findings regarding prediction of persistence versus remission have been inconsistent (Agnew-Blais et al., 2016, 2021; Brinksma et al., 2023; Y.-S. Lee et al., 2025; Riglin et al., 2016). Cognitive functioning has shown more consistent associations: poorer response inhibition, working memory, and reaction-time variability have predicted persistent or worsening symptoms, while improvements in working memory have been associated with ADHD symptom improvement (Karalunas et al., 2017; van Lieshout et al., 2017; Sjöwall, Bohlin, Rydell, & Thorell, 2017; Sudre et al., 2021).

Temperament, particularly negative affect and irritability, may represent an important additional dimension to consider alongside core ADHD symptoms when characterizing developmental trajectories. Irritability and negative affect are elevated among many youth with ADHD and have been associated with greater risk for subsequent depression, anxiety, ODD, and suicidality (Eyre et al., 2017, 2019; Karalunas, Gustafsson, Fair, Musser, & Nigg, 2019; Silva Santos et al., 2025; Vidal-Ribas, Brotman, Valdivieso, Leibenluft, & Stringaris, 2016). Moreover, irritability shows meaningful stability over time and distinguishes youth with ADHD at greater risk for later impairment and psychiatric comorbidity (Karalunas et al., 2019). Thus, modeling the longitudinal course of irritability in parallel with inattention and hyperactivity/impulsivity may capture clinically meaningful heterogeneity not reflected by core ADHD symptoms alone and provide additional information about risk for later comorbid outcomes.

### Heterogeneity in outcomes

Youth with ADHD are at elevated risk for adverse outcomes, including substance use, educational and social difficulties, psychiatric comorbidity, and suicidality (Balazs & Kereszteny, 2017; Erskine et al., 2016; S. S. Lee, Humphreys, Flory, Liu, & Glass, 2011; Septier, Stordeur, Zhang, Delorme, & Cortese, 2019). Emerging evidence suggests that this risk varies according to symptom course. Persistently elevated ADHD symptoms have been associated with greater psychological distress, social isolation, aggression, and poorer educational attainment (Sasser et al., 2016), while late-onset symptoms have also been linked to poorer psychosocial outcomes (Carter, Speyer, Caye, Rohde, & Murray, 2025). However, associations between individual differences in symptom change and outcomes such as substance use and new psychiatric disorder onset remain poorly understood. Clarifying these associations may improve identification of youth at greatest risk for adverse outcomes.

### Current study

The present study addressed prior gaps by examining individual-level trajectories of inattention, hyperactivity/impulsivity, and irritability across development. We examined a broad set of prospective predictors—including polygenic risk, early life environmental risk factors, and cognition—and evaluated whether individual differences in symptom trajectories predicted new psychiatric disorder onset, substance use, and suicidality. Analyses were conducted in two large longitudinal cohorts, Oregon ADHD-1000 and the Adolescent Brain Cognitive Development (ABCD) Study. Given differences in ascertainment, age, and measurement across cohorts, comparisons were used to evaluate the generalizability and robustness of findings rather than as direct replication tests.

## MATERIALS AND METHODS

### Participants

The Oregon-ADHD-1000 cohort consists of a community recruited case-control sample of children aged 7 to 11 years at baseline and followed for up to 12 years (Nigg et al., 2023). Analyses were performed with data from a total of N=1215 (ADHD N=598) participants. Approval for the use of human participants and ethics considerations were obtained from the local University Institutional Review Board. Written informed consent was obtained from the parents of children participating in the Oregon-ADHD-1000 study. Children provided written assent.

The ABCD study cohort is a 21-site, nationwide collected sample of children aged 9 to 10 years at baseline and followed longitudinally. This data is publicly available on the NIMH Data Archive (Agnew-Blais et al., 2016; Barch et al., 2021; Carter et al., 2025, p. 2; Uban et al., 2018). The ABCD study data release 5.1, which includes data for the baseline visit (ages 9-10 at baseline) and four annual follow-up visits, was used for the current study (10.15154/z563-zd24). The participating children were genotyped and evaluated using an extensive and broad range of measures (including imaging, cognitive tasks, and symptom surveys) on an annual basis. For the current study, analyses were performed on a matched ADHD case-control subsample from the larger ABCD study (N=1252, ADHD N=418), which has been described previously (Mooney et al., 2023, 2025). See Table 1 for an overview of the two samples and Supplemental Materials for further details.

**Table 1.** Descriptive statistics for subsets of participants in the Oregon ADHD-1000 and ABCD cohort at study baseline. The ABCD Case-Control subsample is a matched ADHD-control sample based on race, ethnicity, sex, and age (see Supplemental Materials). Inattention and hyperactivity are measured using mean KSADS composite scores in the ABCD cohort (Fan et al., 2021), and ADHD-RS raw scores in the Oregon ADHD-1000 (Nigg et al., 2023).

|  | ABCD Case-Control Subsample | Oregon ADHD-1000 |
| --- | --- | --- |
| Total *N* | 1,252 | 1,215 |
| Mean age (in years) at study year 0 (SD) | 9.17 (0.16) | 7.84 (0.43) |
| % Female | 31.63% | 39.92% |
| % White, European | 50.48% | 79.92% |
| % Hispanic, Latino | 18.69% | 5.02% |
| % Black, African-American | 16.69% | 6.42% |
| % ADHD Cases | 33.3% | 49.17% |
| Mean inattention symptoms (SD) | 0.28 (0.40) | 10.66 (7.95) |
| Mean hyperactivity symptoms (SD) | 0.24 (0.36) | 9.40 (7.70) |
| History of lifetime ADHD medication usage | 9.85% | 34.04% |

### Measures of ADHD Symptoms

Primary analyses focused on measures of inattention, hyperactivity, and irritability deemed most comparable across the two cohorts. In the Oregon-ADHD-1000 cohort, inattention and hyperactivity were measured with raw scores from the parent-reported ADHD Rating Scale (ADHDRS). In addition, irritability was measured using a sum score derived from three items (often loses temper; often touchy or easily annoyed; often angry or resentful) from the Kiddie Schedule for Affective Disorders and Schizophrenia (KSADS). Each item was coded as present/absent resulting in a sum score ranging from 0 to 3.

In the ABCD cohort, inattention and hyperactivity were measured using the parent-reported computerized KSADS (KSADS-COMP; note: for the Oregon ADHD-1000, KSADS were clinician-administered, hence the decision to use the ADHDRS measures). Composite scores for each domain were derived by averaging 9 relevant items as described previously (Cordova et al., 2022). Irritability was measured as a sum of three items from the Child Behavior Checklist (CBCL) (Achenbach, Dumenci, & Rescorla, 2003) whose content was similar (temper tantrums or hot temper; sudden changes in mood or feelings; stubborn, sullen, or irritable) to that of the KSADS items used in the Oregon ADHD-1000. However, the CBCL items were ordinal (0=not true, 1=somewhat or sometimes true, 2=very true), yielding a sum score ranging from 0 to 6. The CBCL measures were chosen to avoid a direct dependency between the irritability measure and the KSADS diagnostic items used to assess new onsets of disruptive disorders (see Adverse Mental Health Outcomes below).

Additional, cohort-specific symptom measures were examined in secondary analyses. See Supplemental Materials for details.

### Early-Childhood Predictors

Several factors hypothesized to explain variation in ADHD trajectories were examined in both cohorts. These early-childhood factors encompassed measures of community-level environment, family and social environment, polygenic risk, and cognition.

### Geocoded exposure measures

In both Oregon ADHD-1000 and ABCD, geocoded estimates of exposures were based on the residential address of each participant. In ABCD, this included the primary address of the participant at the baseline study visit (ages 9-10 years). Exposure estimates based on these geocodes included the childhood opportunity index (COI; total score and three subscores: education, health/environment, and social/economic; https://www.diversitydatakids.org/child-opportunity-index), the area deprivation index (ADI) (Singh, 2003), lead exposure risk (Fan et al., 2021), NO_2_ exposure (Di et al., 2020, no. 2), and particulate matter (PM2.5) exposure (Di et al., 2019).

### Family and social environment measures

In the Oregon ADHD-1000, family conflict, parent stress or trauma, and parenting measures were assessed using the Family Environment Scale (FES) (Moos & Moos, 2002) at baseline, Perceived Stress Scale (PSS) (Cohen, Kamarck, & Mermelstein, 1983) at the 4-year follow-up, and the Alabama Parenting Questionnaire (Frick, 1991) (positive parenting, involvement, poor monitoring, and inconsistent discipline subscales) at the 1-year follow-up.

In ABCD, family conflict and parental stress were also measured using the FES (Moos & Moos, 2002) (at baseline) and PSS (Cohen et al., 1983) (at the 3-year follow-up). For baseline parenting behaviors, positive parenting/emotional warmth was measured using the Child’s Report of Parent Behavior Inventory (CRPBI) (Schaefer, 1965) and parental monitoring was measured using the youth-reported Parental Monitoring Questionnaire (PMQ) (Karoly, Callahan, Schmiege, & Feldstein Ewing, 2016). In addition, educational support and neglectful behavior using the youth-reported Multidimensional Neglectful Behavior Scale (Dubowitz et al., 2011) (MNBS; educational support mean and total mean scores) were collected at the 4-year follow-up visit.

### Polygenic risk

In both cohorts, ADHD polygenic risk scores (PRS) were calculated using PRS-CSx (Ruan et al., 2022) as described previously (Mooney et al., 2025).

### Cognitive measures

In the Oregon ADHD-1000, high-level composite measures were created from multiple tasks to capture cognitive processing speed, working memory, and arousal using previously reported methods (Nigg et al., 2018). In addition, stop-signal reaction time (SSRT) was used to measure response inhibition. In ABCD, cognitive processing speed was measured using the NIH Toolbox Pattern Comparison Processing Speed Test, working memory was measured using the NIH Toolbox List Sorting Working Memory Test, and response inhibition was measured using both the Flanker task and SSRT (Logan & Cowan, 1984; Weintraub et al., 2014).

### Other risk factors

Within Oregon ADHD-1000, the following covariates were considered confounding risk factors (Baker et al., 2023; Willoughby et al., 2020) which were controlled for in our sensitivity analyses: child birth weight in ounces, gestational age measured as the number of weeks premature, the age of the biological mother at the time the child was born, highest household income achieved, highest household employment achieved, highest household education achieved, and the ADHD status of the biological mother.

Within ABCD, the considered risk factors were kept as consistent as possible and also included birth weight, gestational age, mother’s age at the time of child’s birth, highest household income achieved, and highest household education achieved; see (Barch et al., 2021) for details on how these were defined and collected. The Adult Self Report ADHD DSM-oriented T-score was used as a proxy for parental ADHD (Achenbach, 2009).

### Adverse Mental Health Outcomes

In both cohorts, six outcomes were examined in relation to ADHD symptom trajectories, including substance use (alcohol and marijuana), suicidality, and the onset of new anxiety (separation anxiety, social phobia, or generalized anxiety disorder), mood (major depressive disorder or dysthymia), and/or disruptive disorders (oppositional defiance disorder or conduct disorder) after the baseline visit. Additionally, the indicators for anxiety, mood, and disruptive disorders were collapsed into a single variable indicating any new disorder onset. In the Oregon ADHD-1000, new disorder onsets were based on clinician consensus diagnoses determined at each annual visit. In the ABCD cohort, new disorder onsets were based on KSADS-COMP (Kaufman, Kobak, Birmaher, & de Lacy, 2021; Townsend et al., 2020) diagnostic items for the same disorders. In ABCD, anxiety disorders also included panic disorder and agoraphobia. Due to sparsity of data at early years, suicidality and substance use were examined only at ages 13 years and older in both cohorts. The individual measures used to define the substance use and suicidality outcomes are described in the Supplemental Materials.

### Statistical Analyses

All analyses were run using Mplus Version 8.10. Data were cleaned and processed using JupyterLab version 4.0.11 and R version 4.3.2. In both the Oregon ADHD-1000 and ABCD samples, the symptom variables were age-binned such that measurements from each study year were assigned to the closest full-year age (e.g., a measurement taken at 8.25 years was assigned to the 8-year age bin). Ages at the half-year mark are rounded to the nearest even number to prevent statistical bias per the integer function in R. Age bins were collapsed due to data sparsity at the extremes of the age distribution. In the Oregon ADHD-1000, ages 7 and 8 were collapsed into a single bin, as were ages 18 through 23. In the ABCD sample, the age range spanned 9 to 16 years, and only the later age bins were collapsed (ages 14 through 16). Supplemental Tables S1 and S2 show the samples sizes in each age bin for the two samples.

Growth models were applied to the age-binned data, and the simplest model that demonstrated satisfactory fit statistics was selected. This entailed checking the CFI and RMSE against AIC and BIC measures of fit with every increase in complexity and ruling out more complex models that demonstrated diminishing returns. Special attention was paid to whether intercept, slope, and quadratic term (where applicable) were statistically significant and simpler models were preferred when those terms were not significant (e.g., a model with a significant fixed quadratic term was selected over a non-significant random quadratic term). We compared linear and quadratic models for every symptom, with combinations of fixed and random variance for the intercept, slope, and quadratic slope term where applicable. BIC was used, along with mean parameter estimate significance and effect size, to assess model complexity (e.g., linear vs. quadratic slopes and fixed vs. random growth parameters). Given the small range of values for the KSADS irritability measure in the Oregon ADHD-1000 cohort, this was modeled as a Poisson-distributed count measure, thus Bayesian information criteria (BIC) and log likelihood (LL) estimates were used to evaluate fit.

Following identification of the best-fitting growth models in each sample, individual-specific growth parameters were exported, and linear regression models were used to test for association between various risk factors and symptom trajectories (i.e., individual-level intercept and linear slope). When testing for associations between risk factors and trajectory slope, models were adjusted for the baseline symptom level (i.e., the individual-level intercept), to account for correlation between trajectory intercept and slope. (Note: we will refer to the baseline symptom level as “intercept” throughout the paper.) Logistic regression models were used to test for association between symptom trajectory parameters and the binary outcome variables. Each test of symptom progression regressed on risk factors, and outcomes regressed on symptoms, were run as individual, discrete models, resulting in 25 tests (8 geocoded, 6 family and social environment, 4 cognitive, and 7 outcomes) per growth model, as each symptom domain was evaluated separately. We therefore used a conservative Bonferroni adjusted p-value threshold defined as 0.05/25=0.002 to determine statistical significance for our results.

When fitting the growth models, missing data on symptom variables was handled in Mplus using full information maximum likelihood (FIML). Likewise, FIML was also used when performing linear regressions examining associations between early-life risk factors and symptom trajectories. In the case of logistic regressions examining associations between symptom trajectories and binary outcome variables, sample sizes for analyses varied and were limited to the subsample with data available for each outcome. A complete overview of variable missingness within each sample is presented in the Supplemental Materials (Tables S1-S5) .

For those predictors and outcomes significantly associated with symptom trajectory parameters, sensitivity analyses were conducted by including covariates for the known ADHD risk factors described above. In addition, effects of sex and history of ADHD medication use were investigated.

## RESULTS

### Growth Models

Growth model fits were excellent across the board in both samples. All models achieved a comparative fit index (CFI) > 0.96, root mean square error of approximation (RMSEA) < 0.04, and standardized root mean square residual (SRMR) < 0.08. All growth model fit statistics are reported in Table S6.

In the Oregon ADHD-1000 sample, linear models with random intercept and slope were generally found to fit best, except for parent-reported hyperactivity, for which the best-fitting model was a quadratic model with the quadratic term variance fixed at 0. In the ABCD subsample, generally, quadratic models with fixed variance on the quadratic term fit best, although quadratic terms were generally small.

For all symptoms, in both cohorts, mean growth parameter estimates indicate decreasing symptoms over time, though the extent varied by domain (Figures S1 and S2). For those with elevated symptoms at intercept, hyperactivity appeared to decrease faster than inattention in both cohorts (see Table 2; Oregon inattention slope = -0.083 (p = 0.007), Oregon hyperactivity slope = -0.658 (p < 0.001); ABCD inattention slope = -0.048 (p < 0.001), ABCD hyperactivity slope = -0.066 (p < 0.001). Irritability showed a consistent decreasing trajectory across the two cohorts. Mean growth parameter estimates for all symptoms are shown in Table 2.

**Table 2.** Growth model parameter estimates. Abbreviations: I = Intercept; S = Slope; Q = Quadratic; SE = Standard Error; ADHDRS =ADHD Rating Scale; KSADS = Kiddie Schedule for Affective Disorders and Schizophrenia; KSADS-COMP = Kiddie Schedule for Affective Disorders and Schizophrenia – Computerized.

| Oregon ADHD-1000 |  |  |  |  |
| --- | --- | --- | --- | --- |
| Symptom | Term | Est. | SE | P-value |
| ADHDRS - Inattention | I | 11.141 | 0.250 | < 0.001 |
|  | S | -0.083 | 0.031 | 0.007 |
| ADHDRS - Hyperactivity | I | 9.591 | 0.257 | < 0.001 |
|  | S | -0.658 | 0.074 | < 0.001 |
|  | Q | 0.018 | 0.007 | 0.008 |
| KSADS - Irritability | I | -2.026 | 0.097 | < 0.001 |
|  | S | -0.127 | 0.023 | < 0.001 |
| ABCD Case-Control Subsample |  |  |  |  |
| Symptom |  | Est. | SE | P-value |
| KSADS-COMP - Inattention | I | 0.303 | 0.014 | < 0.001 |
|  | S | -0.048 | 0.008 | < 0.001 |
|  | Q | 0.006 | 0.001 | < 0.001 |
| KSADS-COMP - Hyperactivity | I | 0.280 | 0.014 | < 0.001 |
|  | S | -0.066 | 0.007 | < 0.001 |
|  | Q | 0.007 | 0.001 | < 0.001 |
| CBCL - Irritability | I | 1.355 | 0.059 | < 0.001 |
|  | S | -0.129 | 0.035 | < 0.001 |
|  | Q | 0.013 | 0.006 | 0.032 |

With the exception of irritability in the Oregon ADHD-1000 cohort, there were significant negative correlations between intercepts and slopes in both samples, indicating that symptoms decreased faster for those with higher intercept symptoms (Oregon ADHD-1000 ADHDRS Inattention intercept/slope (I/S) covariance = -1.96, p <0.001, Pearson correlation = -0.46; Oregon ADHD-1000 ADHDRS Hyperactivity I/S covariance = -3.92, p <0.001, Pearson correlation = -0.51; Oregon ADHD-1000 KSADS irritability I/S covariance = 0.034, p = 0.10, Pearson correlation = 0.19; ABCD KSADS-COMP Inattention I/S covariance = -0.009, p <0.001, Pearson correlation = -0.78; ABCD KSADS-COMP Hyperactivity I/S covariance = -0.014, p <0.001, Pearson correlation = -0.89, ABCD CBCL Irritability I/S covariance = -0.141, p <0.001, Pearson correlation = -0.59). In our regression analyses, when examining associations with symptom slope, we control for individual-level intercept to rule out indirect effects (i.e., associations simply due to the correlation between intercept and slope), which is justified based on the significant correlations observed.

### Symptom Trajectory Associations

#### Intercept Symptoms

##### Geocoded exposure measures

No significant associations with intercept symptom levels were observed for any community environment variables in either cohort after multiple testing correction (Table S7.

##### Family/social environment measures and polygenic risk

Measures of family conflict and parental stress were significantly and positively associated with intercept levels for all symptoms in both cohorts. In addition, intercept-level inattention was significantly associated with poor parental monitoring in both cohorts.

For hyperactivity and irritability at intercept, similar associations with adverse parenting styles were observed, though the specific measures differed across the two samples (Figure 1 and Table S8).

**Figure 1.**
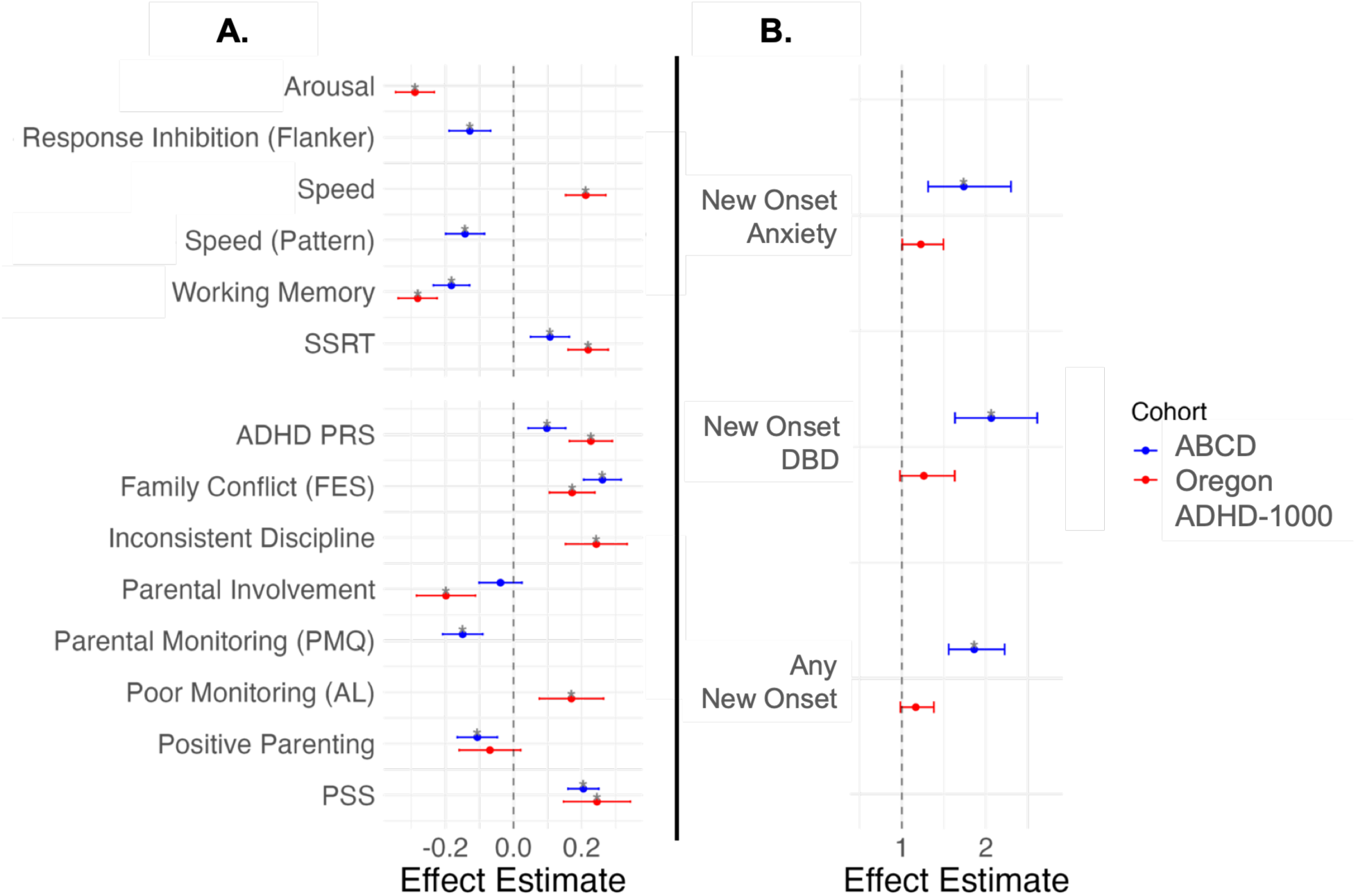
Significant associations with intercept inattention in the ABCD Case-Control subsample and Oregon ADHD-1000 cohorts. (A) Effect estimates for early-life risk factors (standardized regression coefficient with standard errors). (B) Logistic regression effect estimates for risk of later outcomes (odds ratio with 95% confidence intervals). An effect estimate was plotted if the effect was significant after multiple-testing correction (p < 0.002; indicated by *) in at least one cohort. Inattention was measured using the ADHD-RS inattention raw score in the Oregon ADHD-1000 cohort and an inattention composite score from the KSADS-COMP in the ABCD Case-Control subsample. SSRT = stop signal reaction time; PRS = polygenic risk score; FES = family environment scale; PMQ = parental monitoring questionnaire; AL = Alabama Parenting Questionnaire; PSS = perceived stress questionnaire; DBD = disruptive behavior disorder).

ADHD PRS was significantly associated with inattention and hyperactivity at intercept (p < 0.001) in both samples. An association between ADHD PRS and intercept irritability was significant in the Oregon ADHD-1000 but did not survive multiple testing correction in the ABCD subsample (Oregon = 0.158, p < 0.001; ABCD = 0.069, p-value=0.016) (Figure 1 and Table S8).

##### Cognitive measures

There were significant associations between cognitive measures and intercept hyperactivity and inattention in both samples (Figure 1 and Table S9). In the Oregon ADHD-1000, higher speed factor scores and higher SSRT (i.e., longer reaction times), as well as lower working memory and arousal factor scores, were predictive of higher symptoms at intercept. In the ABCD subsample, higher intercept symptom levels were associated with poorer performance on each of the three cognitive tasks examined. Associations between irritability and cognition (poorer performance on the list sorting task and increased SSRT) were seen only in the ABCD subsample following multiple testing corrections, although the KSADS irritability was associated with working memory in the Oregon cohort before correction (uncorrected p-value=0.033).

##### Adverse mental health outcomes

Associations between intercept symptom levels and outcomes were mixed between the two samples. In the Oregon ADHD-1000, only intercept irritability was associated with new disorder onsets, though these did not survive multiple testing correction (Figure 1 and Table S10). In the ABCD subsample, higher intercept levels for all symptoms were associated with increased risk of new disorder onsets (Table S10).

In the Oregon ADHD-1000 alone, intercept irritability was significantly associated with higher risk for suicidality (Table S10).

#### Symptom Change Over Time

There were no generalizable associations between symptom variation over time (slope) and any of the early-life risk factors examined after adjusting for intercept symptom level (Tables S11-S13). In the Oregon ADHD-1000 alone, worse performance on both working memory and speed factors were predictive of a more positive slope for inattention (i.e., a worsening or slower decline of symptoms over time; p < 0.001) (Table S13).

However, in both samples, an increased probability of new onsets of disruptive disorders was positively associated with the slope of irritability. A similar trend was observed for inattention and hyperactivity, though those associations did not survive multiple testing correction (Figure 2 and Table 3).

**Figure 2.**
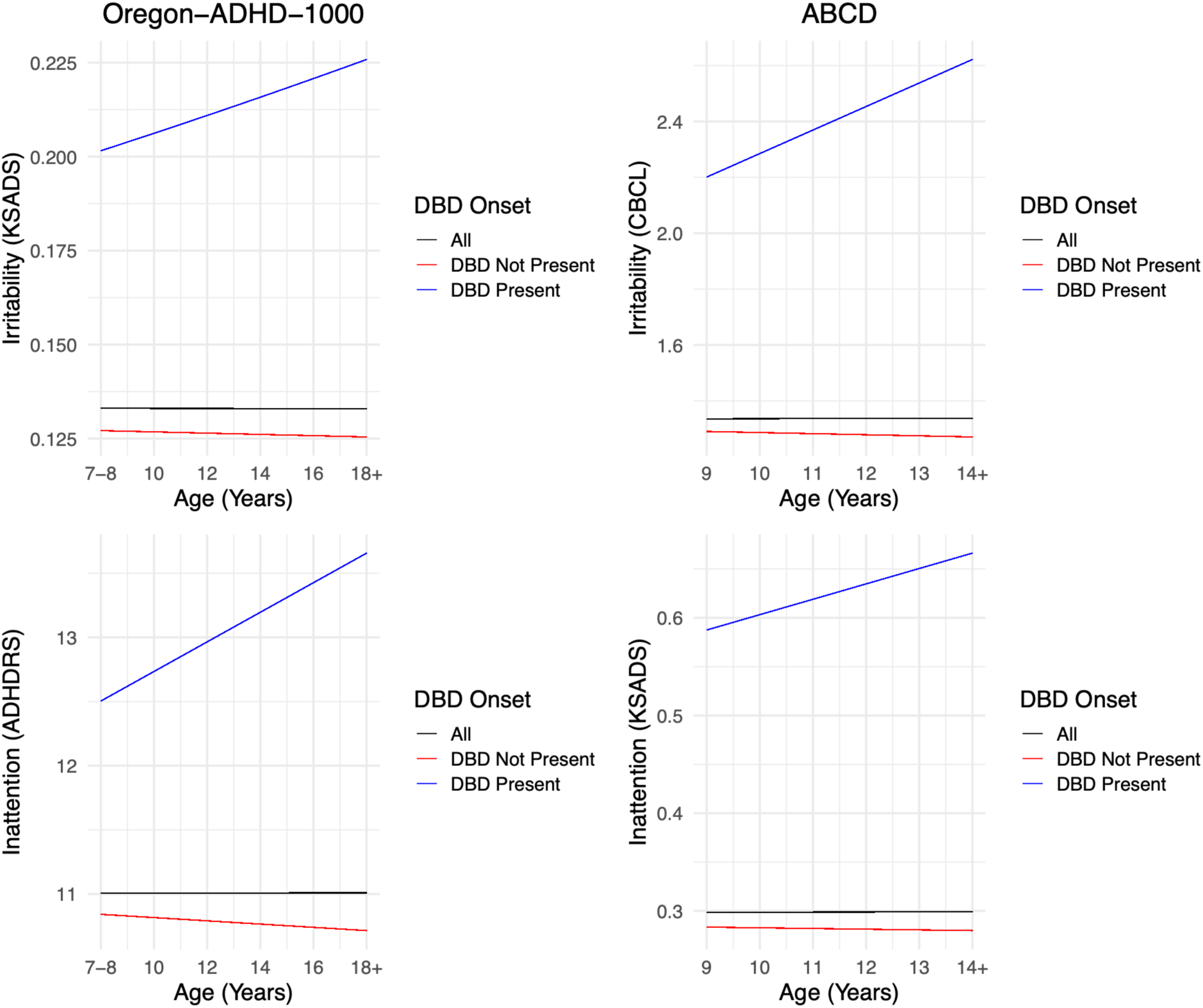
Disruptive behavior disorder onset against ADHD symptom trajectory with residualized linear slope. Top left: Oregon-ADHD-1000, KSADS irritability. Bottom left: Oregon-ADHD-1000, ADHDRS inattention. Note that the effect for disruptive disorder was nominally significant for Oregon-ADHD-1000 inattention (p < 0.007). Top right: ABCD Case-Control subsample, CBCL irritability. Bottom right: ABCD Case-Control subsample, KSADS-COMP inattention. Because the variance was fixed on the quadratic term in the ABCD models, we have excluded the q-term from the trajectory plots for clarity.

**Table 3.**
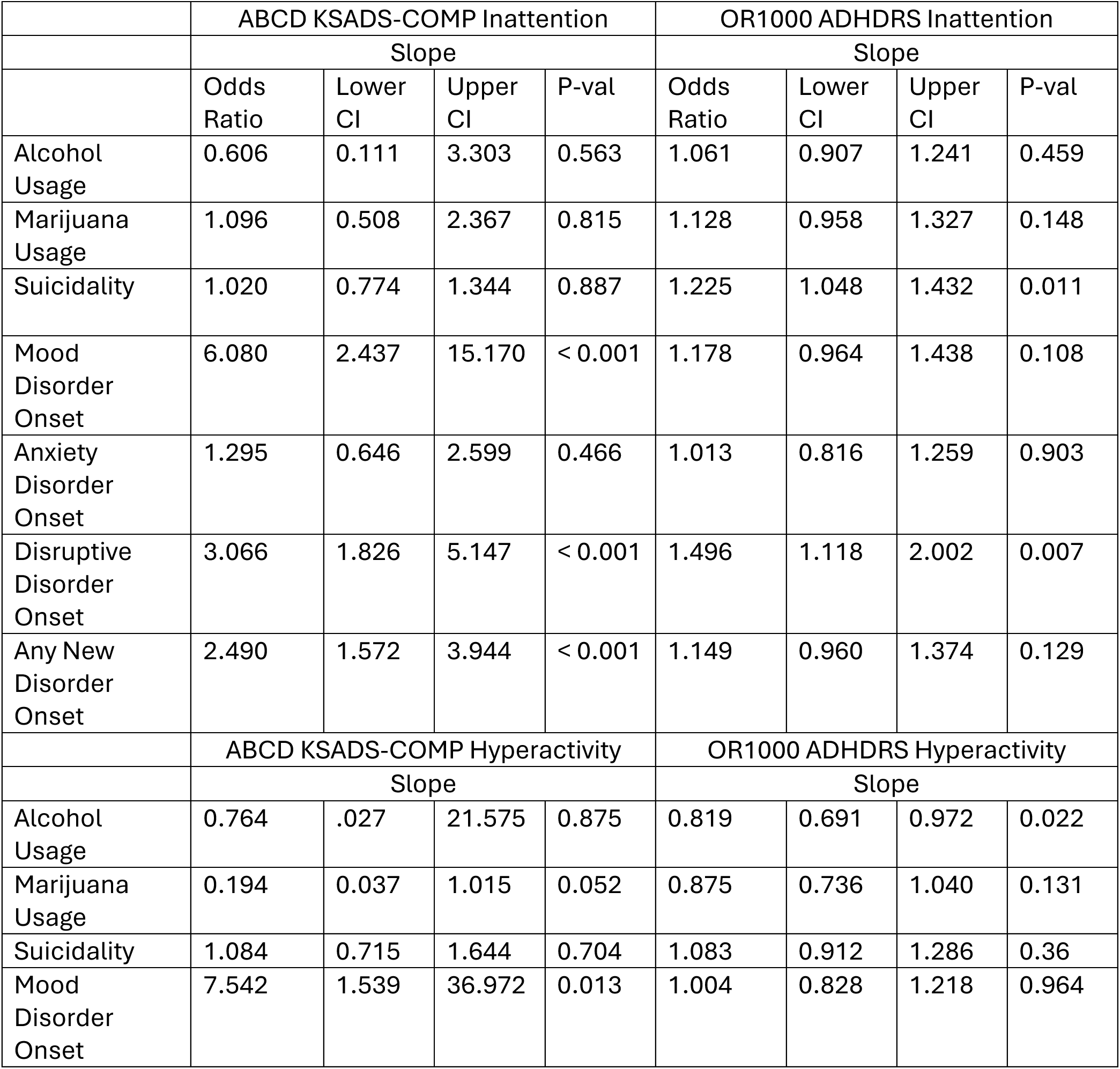
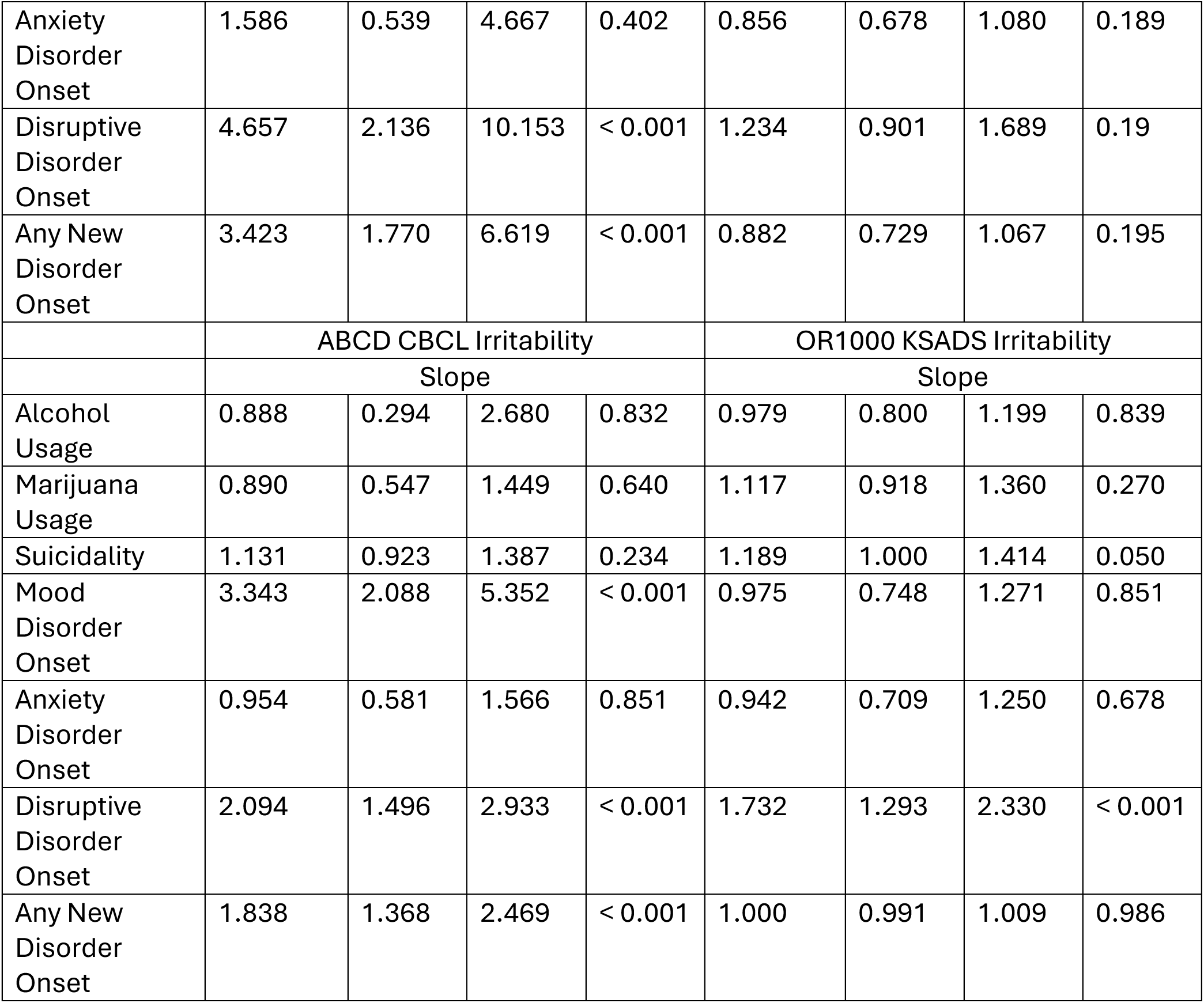
Associations between trajectory (slope) ADHD inattention, hyperactivity, and irritability symptoms in the ABCD Case-Control subsample and Oregon ADHD-1000 cohorts and behavioral outcomes. Abbreviations: Est. = Regression standardized; Coefficient, SE = standard error; CI = confidence interval (95%?)

|  | ABCD KSADS-COMP Inattention |  |  |  | OR1000 ADHDRS Inattention |  |  |  |
| --- | --- | --- | --- | --- | --- | --- | --- | --- |
|  | Slope |  |  |  | Slope |  |  |  |
|  | Odds Ratio | Lower CI | Upper CI | P-val | Odds Ratio | Lower CI | Upper CI | P-val |
| Alcohol Usage | 0.606 | 0.111 | 3.303 | 0.563 | 1.061 | 0.907 | 1.241 | 0.459 |
| Marijuana Usage | 1.096 | 0.508 | 2.367 | 0.815 | 1.128 | 0.958 | 1.327 | 0.148 |
| Suicidality | 1.020 | 0.774 | 1.344 | 0.887 | 1.225 | 1.048 | 1.432 | 0.011 |
| Mood Disorder Onset | 6.080 | 2.437 | 15.170 | < 0.001 | 1.178 | 0.964 | 1.438 | 0.108 |
| Anxiety Disorder Onset | 1.295 | 0.646 | 2.599 | 0.466 | 1.013 | 0.816 | 1.259 | 0.903 |
| Disruptive Disorder Onset | 3.066 | 1.826 | 5.147 | < 0.001 | 1.496 | 1.118 | 2.002 | 0.007 |
| Any New Disorder Onset | 2.490 | 1.572 | 3.944 | < 0.001 | 1.149 | 0.960 | 1.374 | 0.129 |
|  | ABCD KSADS-COMP Hyperactivity |  |  |  | OR1000 ADHDRS Hyperactivity |  |  |  |
|  | Slope |  |  |  | Slope |  |  |  |
|  | Odds Ratio | Lower CI | Upper CI | P-val | Odds Ratio | Lower CI | Upper CI | P-val |
| Alcohol Usage | 0.764 | .027 | 21.575 | 0.875 | 0.819 | 0.691 | 0.972 | 0.022 |
| Marijuana Usage | 0.194 | 0.037 | 1.015 | 0.052 | 0.875 | 0.736 | 1.040 | 0.131 |
| Suicidality | 1.084 | 0.715 | 1.644 | 0.704 | 1.083 | 0.912 | 1.286 | 0.36 |
| Mood Disorder Onset | 7.542 | 1.539 | 36.972 | 0.013 | 1.004 | 0.828 | 1.218 | 0.964 |
| Anxiety Disorder Onset | 1.586 | 0.539 | 4.667 | 0.402 | 0.856 | 0.678 | 1.080 | 0.189 |
| Disruptive Disorder Onset | 4.657 | 2.136 | 10.153 | < 0.001 | 1.234 | 0.901 | 1.689 | 0.19 |
| Any New Disorder Onset | 3.423 | 1.770 | 6.619 | < 0.001 | 0.882 | 0.729 | 1.067 | 0.195 |
|  | ABCD CBCL Irritability |  |  |  | OR1000 KSADS Irritability |  |  |  |
|  | Slope |  |  |  | Slope |  |  |  |
| Alcohol Usage | 0.888 | 0.294 | 2.680 | 0.832 | 0.979 | 0.800 | 1.199 | 0.839 |
| Marijuana Usage | 0.890 | 0.547 | 1.449 | 0.640 | 1.117 | 0.918 | 1.360 | 0.270 |
| Suicidality | 1.131 | 0.923 | 1.387 | 0.234 | 1.189 | 1.000 | 1.414 | 0.050 |
| Mood Disorder Onset | 3.343 | 2.088 | 5.352 | < 0.001 | 0.975 | 0.748 | 1.271 | 0.851 |
| Anxiety Disorder Onset | 0.954 | 0.581 | 1.566 | 0.851 | 0.942 | 0.709 | 1.250 | 0.678 |
| Disruptive Disorder Onset | 2.094 | 1.496 | 2.933 | < 0.001 | 1.732 | 1.293 | 2.330 | < 0.001 |
| Any New Disorder Onset | 1.838 | 1.368 | 2.469 | < 0.001 | 1.000 | 0.991 | 1.009 | 0.986 |

#### Sensitivity Analyses

Sensitivity analyses were conducted to adjust for a set of potentially confounding risk factors. Lower maternal age at birth and maternal ADHD status were significantly associated with higher intercept levels for all symptoms in both cohorts (all p-values <= 0.001). Likewise, in both cohorts, lower household income was also associated with higher levels of all symptoms, though the association with inattention did not survive multiple testing correction in the ABCD subsample (Table S14).

In the ABCD subsample alone, lower birth weight, maternal age at birth, household income, and household education level were significantly associated with more positive symptom slope (i.e., more stable symptoms over time; Table S15).

Adjusting for these covariates (see Methods) had minimal impact on the robust associations identified in our primary analyses (reported above). In the Oregon ADHD-1000, the associations between intercept irritability and parental stress as well as parental involvement did not survive multiple testing correction in the covariate-adjusted models (though parental involvement, at p = 0.003, can be considered nominally significant). And in the ABCD subsample, associations between intercept hyperactivity and performance on both the Flanker and pattern comparison processing speed tasks were no longer significant. All other associations that generalized across the two cohorts, as well as the associations between cognition and symptom slope seen in the Oregon ADHD-1000 cohort, remained significant in covariate-adjusted models (Tables S16 and S17).

#### Note About Medication Status

Lifetime medication status (an indicator of whether a child had ever been medicated for ADHD) was significantly associated with all symptoms at intercept (Table S18). In the Oregon ADHD-1000, history of medication use was also significantly associated with inattention and hyperactivity trajectories (slopes) (p-values <0.001), indicating that children with persistent elevated ADHD symptoms were more likely to be medicated. This effect was not seen in the ABCD subsample (Table S19).

## DISCUSSION

Growth models showed an overall decline in symptoms over time, with hyperactivity declining the fastest, and inattention demonstrating a more stable trajectory (Barch et al., 2021; Brinksma et al., 2023; Carter et al., 2025; Durdurak, Morales-Muñoz, Hosang, & Marwaha, 2025; Y.-S. Lee et al., 2025; Sasser et al., 2016; Wootton et al., 2022). Individual trajectories showed a large degree of variability (Figures S1 and S2), which lends credence to the utility of individual-level symptom trajectories when examining relationships to risk factors and adverse outcomes, as group-level analysis may obfuscate important individual differences.

Polygenic risk, early risk factors, and cognition all predicted ADHD symptom severity at the initial assessment in both cohorts, consistent with prior research (Brinksma et al., 2023; Carter et al., 2025; Y.-S. Lee et al., 2025). Effect sizes were generally small to medium. However, none of the early-life risk factors examine here robustly predicted symptom change over time. Cognition significantly predicted inattention symptom change over time within the Oregon ADHD-1000, consistent with prior findings (Karalunas et al., 2017; van Lieshout et al., 2017; Sjöwall et al., 2017; Sudre et al., 2021), but these associations did not generalize to the ABCD cohort. Overall, these findings suggest that the factors distinguishing ADHD cases from controls are not necessarily the same factors that explain heterogeneity in developmental trajectories.

Increasing inattention and irritability symptom trajectories were linked to increased risk for onset of new disorders, particularly mood and disruptive disorders, consistent with prior work (Durdurak et al., 2025; Y.-S. Lee et al., 2025; Sasser et al., 2016). However, ADHD symptom trajectories showed minimal relation to both substance use and suicide outcomes. This and other studies may be limited to detect effects due to the low frequency of usage in each cohort (15.9% in Oregon ADHD-1000; 2.1% in ABCD). Notably, increasing irritability trajectories was associated with increased risk for suicidality in the Oregon-ADHD-1000 cohort only, highlighting the potential relevance of co-occurring irritability symptoms among youth with diagnosed ADHD in childhood for understanding later adverse outcomes, even those with lower base rates.

Findings from the current study should be interpreted with the following limitations in mind. First, differences across cohorts in ascertainment, measurement, and age may have contributed to lack of generalization of findings. For instance, statistically significant associations between measures of cognition and symptom slopes were seen in the Oregon ADHD-1000, but not in the ABCD cohort. Conversely, a significant association between inattention slope and onset of mood disorders was observed in the ABCD cohort alone. The Oregon ADHD-1000 is a clinically ascertained sample enriched for youth with ADHD and greater symptom severity, providing increased variability in cognitive impairment and symptom trajectories that may facilitate detection of cognition–trajectory associations. In contrast, the population-based ABCD cohort includes a broader spectrum of symptom severity and psychopathology, potentially making it better suited to detecting associations with later-emerging outcomes, such as mood disorders.

Second, although this study was conducted within fairly large cohorts, data was sparse at the extremes of the age range (14+ in ABCD and 18+ in Oregon-ADHD-1000). This almost certainly affected the prevalence of the outcomes examined, such as substance use and suicidality, particularly in the ABCD cohort. Given that suicidal behavior often peaks in late adolescence and into early adulthood, it’s not surprising that effects were found in the Oregon-ADHD-1000 and not in ABCD. Follow-up as new waves of data as youth develop into adulthood along with greater measurement granularity will be important. Notably, because we also examined all new disorder onsets, we are unable to make definitive claims about the temporal relationships between symptom trajectories and outcomes.

Finally, most participants in both cohorts are male (approximately 70%). These findings therefore cannot address more nuanced questions regarding how ADHD symptoms evolve over time differently depending on sex. Generalizability across race, ethnicity, and culture also requires more work. While the majority of youth in both cohorts were White and Non-Hispanic, the diversity of the ABCD cohort helps to overcome this somewhat (Barch et al., 2021; Cordova et al., 2022).

Future work may benefit from considering other timescales of change. These data focused on change over years, but there is increasing evidence that ADHD symptoms vary over moments, days, weeks, and months (Ben-Dor Cohen, Maeir, Eldar, & Nahum, 2023; Karalunas, Dude, Figuracion, & Lane, 2024; Kennedy, Molina, & Pedersen, 2024). Future studies might benefit from examining symptom change across multiple timescales in order to connect symptom change more meaningfully to long-term outcomes. Given the heterogeneous nature and presentation of the disorder, identifying the right timescales of symptom change for analysis may be critical for intervention.

Overall, the present study helps to elucidate connections between early risk factors, ADHD symptom trajectories, and potential adverse outcomes over time in a diverse set of participants from two independent cohorts. The results highlight the role of family dynamics, social environment, and cognition on ADHD symptoms and how the course of these symptoms influence risk for adverse outcomes.

## Data Availability

All data produced in the present study are available upon reasonable request to the authors.

https://abcdstudy.org

http://dx.doi.org/10.15154/z563-zd24

## ACKNOWLEDGEMENTS

Data used in the preparation of this article were obtained from the Adolescent Brain Cognitive DevelopmentSM (ABCD) Study (https://abcdstudy.org), held in the NIMH Data Archive (NDA). This is a multisite, longitudinal study designed to recruit more than 10,000 children age 9-10 and follow them over 10 years into early adulthood. The ABCD Study® is supported by the National Institutes of Health and additional federal partners under award numbers U01DA041048, U01DA050989, U01DA051016, U01DA041022, U01DA051018, U01DA051037, U01DA050987, U01DA041174, U01DA041106, U01DA041117, U01DA041028, U01DA041134, U01DA050988, U01DA051039, U01DA041156, U01DA041025, U01DA041120, U01DA051038, U01DA041148, U01DA041093, U01DA041089, U24DA041123, U24DA041147. A full list of supporters is available at https://abcdstudy.org/federal-partners.html. A listing of participating sites and a complete listing of the study investigators can be found at https://abcdstudy.org/consortium_members/. ABCD consortium investigators designed and implemented the study and/or provided data but did not necessarily participate in the analysis or writing of this report. This manuscript reflects the views of the authors and may not reflect the opinions or views of the NIH or ABCD consortium investigators. The ABCD data repository grows and changes over time. The ABCD data used in this report came from the ABCD 5.1 data release (http://dx.doi.org/10.15154/z563-zd24). Additional support for this work was made possible from NIEHS R01-ES032295 and R01-ES031074.

This work was supported by the National Institute of Mental Health of the National Institutes of Health under award number R01MH131685. The project was also supported by the National Center for Advancing Translational Sciences (NCATS), National Institutes of Health, through grant award number UL1TR002369. The content is solely the responsibility of the authors and does not necessarily represent the official views of the National Institutes of Health.

## ETHICS STATEMENT

For the Oregon ADHD-1000 cohort, human subjects and ethics approval was obtained from the Oregon Health & Science University Institutional Review Board under IRB Study #4817 (11/06/2008). A parent/legal guardian provided written informed consent, and children provided written assent for participation in the study.

For the ABCD study cohort, data was accessed through the NIMH Data Archive, which ensures data submitted to the repository is done so in accordance with informed consent of research participants and/or approval from an institutional review board. The data was used in accordance with the NIMH Data Archive Data Use Certification.

## AI STATEMENT

The authors declare that they have not used AI in any capacity that would require disclosure in accordance with Wiley’s AI Guidelines, nor in any capacity that would reasonably require discloser for editors, reviewers and readers to properly evaluate their research or manuscript. The authors confirm that AI has not been used for purposes including but NOT limited to drafting and editing, to generate substantial text or restructure arguments, nor in the research methodology. The authors take full responsibility for the accuracy of this statement.

## BIBLIOGRAPHY

Achenbach, T. M. (2009). Some needed changes in DSM-V: But what about children? Wiley-Blackwell Publishing Ltd.

Achenbach, T. M., Dumenci, L., & Rescorla, L. A. (2003). DSM-Oriented and Empirically Based Approaches to Constructing Scales From the Same Item Pools. Journal of Clinical Child & Adolescent Psychology, 32(3), 328–340. Routledge.

Agnew-Blais, J. C., Belsky, D. W., Caspi, A., Danese, A., Moffitt, T. E., Polanczyk, G. V., Sugden, K., et al. (2021). Polygenic Risk and the Course of Attention-Deficit/Hyperactivity Disorder From Childhood to Young Adulthood: Findings From a Nationally Representative Cohort. Journal of the American Academy of Child & Adolescent Psychiatry, 60(9), 1147–1156.

Agnew-Blais, J. C., Polanczyk, G. V., Danese, A., Wertz, J., Moffitt, T. E., & Arseneault, L. (2016). Evaluation of the Persistence, Remission, and Emergence of Attention-Deficit/Hyperactivity Disorder in Young Adulthood. JAMA psychiatry, 73(7), 713–720. United States.

Agnew-Blais, J. C., Polanczyk, G. V., Danese, A., Wertz, J., Moffitt, T. E., & Arseneault, L. (2020). Are changes in ADHD course reflected in differences in IQ and executive functioning from childhood to young adulthood? Psychological Medicine, 50(16), 2799–2808.

Baker, B. H., Joo, Y. Y., Park, J., Cha, J., Baccarelli, A. A., & Posner, J. (2023). Maternal age at birth and child attention-deficit hyperactivity disorder: Causal association or familial confounding? Journal of child psychology and psychiatry, and allied disciplines, 64(2), 299–310. England.

Balazs, J., & Kereszteny, A. (2017). Attention-deficit/hyperactivity disorder and suicide: A systematic review. World journal of psychiatry, 7(1), 44.

Barch, D. M., Albaugh, M. D., Baskin-Sommers, A., Bryant, B. E., Clark, D. B., Dick, A. S., Feczko, E., et al. (2021). Demographic and mental health assessments in the adolescent brain and cognitive development study: Updates and age-related trajectories. Developmental Cognitive Neuroscience, 52, 101031.

Ben-Dor Cohen, M., Maeir, A., Eldar, E., & Nahum, M. (2023). Everyday Cognitive Control and Emotion Dysregulation in Young Adults With and Without ADHD: An Ecological Momentary Assessment Study. Journal of attention disorders, 27(5), 539–553. United States.

Biederman, J., Mick, E., & Faraone, S. V. (2000). Age-dependent decline of symptoms of attention deficit hyperactivity disorder: Impact of remission definition and symptom type. The American journal of psychiatry, 157(5), 816–818. United States.

Brinksma, D. M., Hoekstra, P. J., de Bildt, A., Buitelaar, J. K., van den Hoofdakker, B. J., Hartman, C. A., & Dietrich, A. (2023). Parental rejection in early adolescence predicts a persistent ADHD symptom trajectory across adolescence. European Child & Adolescent Psychiatry, 32(1), 139–153.

Carter, L., Speyer, L., Caye, A., Rohde, L., & Murray, A. L. (2025). Late adolescent outcomes of different developmental trajectories of ADHD symptoms in a large longitudinal study. European Child & Adolescent Psychiatry, 34(2), 709–719.

Cohen, S., Kamarck, T., & Mermelstein, R. (1983). A Global Measure of Perceived Stress. Journal of Health and Social Behavior, 24(4), 385–396. [American Sociological Association, Sage Publications, Inc.].

Cordova, M. M., Antovich, D. M., Ryabinin, P., Neighbor, C., Mooney, M. A., Dieckmann, N. F., Miranda-Dominguez, O., et al. (2022). Attention-Deficit/Hyperactivity Disorder: Restricted Phenotypes Prevalence, Comorbidity, and Polygenic Risk Sensitivity in the ABCD Baseline Cohort. Journal of the American Academy of Child and Adolescent Psychiatry, 61(10), 1273–1284. United States.

Cortese, S., Bellgrove, M. A., Brikell, I., Franke, B., Goodman, D. W., Hartman, C. A., Larsson, H., et al. (2025). Attention-deficit/hyperactivity disorder (ADHD) in adults: Evidence base, uncertainties and controversies. World psychiatry: Official journal of the World Psychiatric Association (WPA), 24(3), 347–371. Italy.

Di, Q., Amini, H., Shi, L., Kloog, I., Silvern, R., Kelly, J., Sabath, M. B., et al. (2019). An ensemble-based model of PM2.5 concentration across the contiguous United States with high spatiotemporal resolution. Environment International, 130, 104909.

Di, Q., Amini, H., Shi, L., Kloog, I., Silvern, R., Kelly, J., Sabath, M. B., et al. (2020). Assessing NO(2) Concentration and Model Uncertainty with High Spatiotemporal Resolution across the Contiguous United States Using Ensemble Model Averaging. Environmental science & technology, 54(3), 1372–1384. United States.

Dubowitz, H., Villodas, M. T., Litrownik, A. J., Pitts, S. C., Hussey, J. M., Thompson, R., Black, M. M., et al. (2011). Psychometric properties of a youth self-report measure of neglectful behavior by parents. Child Abuse & Neglect, 35(6), 414–424.

Durdurak, B. B., Morales-Muñoz, I., Hosang, G. M., & Marwaha, S. (2025). ADHD symptom trajectories across childhood and early adolescence and risk for hypomanic symptoms in young adulthood. European Psychiatry, 68(1), e37.

Erskine, H. E., Norman, R. E., Ferrari, A. J., Chan, G. C. K., Copeland, W. E., Whiteford, H. A., & Scott, J. G. (2016). Long-Term Outcomes of Attention-Deficit/Hyperactivity Disorder and Conduct Disorder: A Systematic Review and Meta-Analysis. Journal of the American Academy of Child and Adolescent Psychiatry, 55(10), 841–850. United States.

Eyre, O., Langley, K., Stringaris, A., Leibenluft, E., Collishaw, S., & Thapar, A. (2017). Irritability in ADHD: Associations with depression liability. Journal of Affective Disorders, 215, 281–287.

Eyre, O., Riglin, L., Leibenluft, E., Stringaris, A., Collishaw, S., & Thapar, A. (2019). Irritability in ADHD: association with later depression symptoms. European child & adolescent psychiatry, 28(10), 1375–1384. Germany.

Fan, C. C., Marshall, A., Smolker, H., Gonzalez, M. R., Tapert, S. F., Barch, D. M., Sowell, E., et al. (2021). Adolescent Brain Cognitive Development (ABCD) study Linked External Data (LED): Protocol and practices for geocoding and assignment of environmental data. Developmental cognitive neuroscience, 52, 101030. Netherlands.

Faraone, S. V., Bellgrove, M. A., Brikell, I., Cortese, S., Hartman, C. A., Hollis, C., Newcorn, J. H., et al. (2024). Attention-deficit/hyperactivity disorder. Nature Reviews Disease Primers, 10(1), 11.

Frick, P. J. (1991). Alabama parenting questionnaire. Journal of Clinical Child Psychology.

Karalunas, S. L., Dude, J., Figuracion, M., & Lane, S. P. (2024). Momentary Dynamics Implicate Emotional Features in the ADHD Phenotype. Research on child and adolescent psychopathology, 52(9), 1343–1356. United States.

Karalunas, S. L., Gustafsson, H. C., Dieckmann, N. F., Tipsord, J., Mitchell, S. H., & Nigg, J. T. (2017). Heterogeneity in development of aspects of working memory predicts longitudinal attention deficit hyperactivity disorder symptom change. Journal of abnormal psychology, 126(6), 774–792. United States.

Karalunas, S. L., Gustafsson, H. C., Fair, D., Musser, E. D., & Nigg, J. T. (2019). Do we need an irritable subtype of ADHD? Replication and extension of a promising temperament profile approach to ADHD subtyping. Psychological assessment, 31(2), 236–247. United States.

Karoly, H. C., Callahan, T., Schmiege, S. J., & Feldstein Ewing, S. W. (2016). Evaluating the Hispanic paradox in the context of adolescent risky sexual behavior: The role of parent monitoring. Journal of pediatric psychology, 41(4), 429–440. Oxford University Press.

Kaufman, J., Kobak, K., Birmaher, B., & de Lacy, N. (2021). KSADS-COMP perspectives on child psychiatric diagnostic assessment and treatment planning. Journal of the American Academy of Child & Adolescent Psychiatry, 60(5), 540–542. Elsevier.

Kennedy, T. M., Molina, B. S. G., & Pedersen, S. L. (2024). Change in Adolescents’ Perceived ADHD Symptoms Across 17 Days of Ecological Momentary Assessment. Journal of clinical child and adolescent psychology: The official journal for the Society of Clinical Child and Adolescent Psychology, American Psychological Association, Division 53, 53(3), 397–412. England.

Lee, S. S., Humphreys, K. L., Flory, K., Liu, R., & Glass, K. (2011). Prospective association of childhood attention-deficit/hyperactivity disorder (ADHD) and substance use and abuse/dependence: A meta-analytic review. Clinical Psychology Review, 31(3), 328–341.

Lee, Y.-S., Sprong, M. E., Shrestha, J., Smeltzer, M. P., & Hollender, H. (2025). Trajectory Analysis for Identifying Classes of Attention Deficit Hyperactivity Disorder (ADHD) in Children of the United States. Retrieved June 10, 2025, from https://clinical-practice-and-epidemiology-in-mental-health.com/VOLUME/20/ELOCATOR/e17450179298863/FULLTEXT/

van Lieshout, M., Luman, M., Twisk, J. W. R., Faraone, S. V., Heslenfeld, D. J., Hartman, C. A., Hoekstra, P. J., et al. (2017). Neurocognitive Predictors of ADHD Outcome: A 6-Year Follow-up Study. Journal of abnormal child psychology, 45(2), 261–272. United States.

Logan, G. D., & Cowan, W. B. (1984). On the ability to inhibit thought and action: A theory of an act of control. Psychological review, 91(3), 295. American Psychological Association.

Luo, Y., Weibman, D., Halperin, J. M., & Li, X. (2019). A Review of Heterogeneity in Attention Deficit/Hyperactivity Disorder (ADHD). Frontiers in Human Neuroscience, 13, 42.

Mooney, M. A., Ryabinin, P., Morton, H., Selah, K., Gonoud, R., Kozlowski, M., Nousen, E., et al. (2023). Joint polygenic and environmental risks for childhood attention-deficit/hyperactivity disorder (ADHD) and ADHD symptom dimensions. JCPP advances, 3(2), e12152. United States.

Mooney, M. A., Ryabinin, P., Nousen, E., Tipsord, J., Dieckmann, N. F., Karalunas, S. L., Herting, M. M., et al. (2025, June 17). Sibling Control Analysis of Perinatal Health and Family Environment Factors Related to Childhood ADHD Symptoms. medRxiv: The preprint server for health sciences. United States.

Moos, R. H., & Moos, B. S. (2002). Family environment scale manual: Development,Applications,Research (3rd ed., p. 131 p.). Center for Health Care Evaluation,Department of Veterans Affairs and Stanford University Medical Centers,Maind Garden. Retrieved from https://cir.nii.ac.jp/crid/1970304959935212064

Nigg, J. T., Gustafsson, H. C., Karalunas, S. L., Ryabinin, P., McWeeney, S. K., Faraone, S. V., Mooney, M. A., et al. (2018). Working Memory and Vigilance as Multivariate Endophenotypes Related to Common Genetic Risk for Attention-Deficit/Hyperactivity Disorder. Journal of the American Academy of Child and Adolescent Psychiatry, 57(3), 175–182. United States.

Nigg, J. T., Karalunas, S. L., Mooney, M. A., Wilmot, B., Nikolas, M. A., Martel, M. M., Tipsord, J., et al. (2023). The Oregon ADHD-1000: A new longitudinal data resource enriched for clinical cases and multiple levels of analysis. Developmental cognitive neuroscience, 60, 101222. Netherlands.

Riglin, L., Collishaw, S., Thapar, A. K., Dalsgaard, S., Langley, K., Smith, G. D., Stergiakouli, E., et al. (2016). Association of Genetic Risk Variants With Attention-Deficit/Hyperactivity Disorder Trajectories in the General Population. JAMA psychiatry, 73(12), 1285–1292.

Ruan, Y., Lin, Y.-F., Feng, Y.-C. A., Chen, C.-Y., Lam, M., Guo, Z., Ahn, Y. M., et al. (2022). Improving polygenic prediction in ancestrally diverse populations. Nature Genetics, 54(5), 573–580.

Sasser, T. R., Kalvin, C. B., & Bierman, K. L. (2016). Developmental Trajectories of Clinically Significant ADHD Symptoms from Grade 3 through 12 in a High-Risk Sample: Predictors and Outcomes. Journal of abnormal psychology, 125(2), 207–219.

Schaefer, E. S. (1965). Children’s reports of parental behavior: An inventory. Child development, 413–424. JSTOR.

Septier, M., Stordeur, C., Zhang, J., Delorme, R., & Cortese, S. (2019). Association between suicidal spectrum behaviors and attention-deficit/hyperactivity disorder: A systematic review and meta-analysis. Neuroscience & Biobehavioral Reviews, 103, 109–118. Elsevier.

Silva Santos, R. M., de Andrade, J. N., Soares da Silva, A., Coelho Martins, T., Martins Rezende, V. H., Maia Gomes, P. G., Alvim-Soares Júnior, A. M., et al. (2025). The Meaning of Irritability in ADHD: Systematic Review. Developmental neuropsychology, 50(3–8), 168–184. England.

Singh, G. K. (2003). Area deprivation and widening inequalities in US mortality, 1969-1998. American journal of public health, 93(7), 1137–1143. United States.

Sjöwall, D., Bohlin, G., Rydell, A.-M., & Thorell, L. B. (2017). Neuropsychological deficits in preschool as predictors of ADHD symptoms and academic achievement in late adolescence. Child neuropsychology: A journal on normal and abnormal development in childhood and adolescence, 23(1), 111–128. England.

Sudre, G., Sharp, W., Kundzicz, P., Bouyssi-Kobar, M., Norman, L., Choudhury, S., & Shaw, P. (2021). Predicting the course of ADHD symptoms through the integration of childhood genomic, neural, and cognitive features. Molecular psychiatry, 26(8), 4046–4054. England.

Thapar, A., & Rice, F. (2021). Family-Based Designs that Disentangle Inherited Factors from Pre- and Postnatal Environmental Exposures: In Vitro Fertilization, Discordant Sibling Pairs, Maternal versus Paternal Comparisons, and Adoption Designs. Cold Spring Harbor perspectives in medicine, 11(3). United States.

Townsend, L., Kobak, K., Kearney, C., Milham, M., Andreotti, C., Escalera, J., Alexander, L., et al. (2020). Development of Three Web-Based Computerized Versions of the Kiddie Schedule for Affective Disorders and Schizophrenia Child Psychiatric Diagnostic Interview: Preliminary Validity Data. Journal of the American Academy of Child & Adolescent Psychiatry, 59(2), 309–325. Elsevier.

Uban, K. A., Horton, M. K., Jacobus, J., Heyser, C., Thompson, W. K., Tapert, S. F., Madden, P. A. F., et al. (2018). Biospecimens and the ABCD study: Rationale, methods of collection, measurement and early data. Developmental cognitive neuroscience, 32, 97–106. Netherlands.

Vidal-Ribas, P., Brotman, M. A., Valdivieso, I., Leibenluft, E., & Stringaris, A. (2016). The Status of Irritability in Psychiatry: A Conceptual and Quantitative Review. Journal of the American Academy of Child & Adolescent Psychiatry, 55(7), 556–570.

Weintraub, S., Dikmen, S. S., Heaton, R. K., Tulsky, D. S., Zelazo, P. D., Slotkin, J., Carlozzi, N. E., et al. (2014). The cognition battery of the NIH toolbox for assessment of neurological and behavioral function: Validation in an adult sample. Journal of the International Neuropsychological Society: JINS, 20(6), 567–578. England.

Willoughby, M. T., Williams, J., Mills-Koonce, W. R., & Blair, C. B. (2020). Early Life Predictors of Attention Deficit/Hyperactivity Disorder Symptomatology Profiles from Early Through Middle Childhood. Development and psychopathology, 32(3), 791–802.

Wootton, R. E., Riglin, L., Blakey, R., Agnew-Blais, J., Caye, A., Cadman, T., Havdahl, A., et al. (2022). Decline in attention-deficit hyperactivity disorder traits over the life course in the general population: Trajectories across five population birth cohorts spanning ages 3 to 45 years. International Journal of Epidemiology, 51(3), 919–930.

