## Supplemental Materials for "Longitudinal trajectories of ADHD symptoms: relationships to early-childhood predictors and later outcomes"

#### Supplemental Methods

*ADHD assessment.*

In the Oregon-ADHD-1000 cohort, symptom ratings were derived from parent and teacher reports using standardized and nationally normed rating scales alongside semi-structured clinical parent interviews administered by a clinically trained interviewer, notes from two trained research assistants, and child observation all with strong inter-interviewer reliability. Two clinicians, one a board-certified child psychiatrist and the other a licensed child clinical psychologist, reviewed the collected data independently to arrive at a diagnosis decision (disagreements decided by consensus) with adequate agreement (k > 0.80). See (Nigg et al., 2018) for further details.

In the ABCD cohort, ADHD status was determined using methods outlined by (Cordova et al., 2022). However, Cordova et al.'s criteria was expanded to include participants who had met the diagnostic criteria for ADHD in the past and continued to display elevated symptoms. This was determined by participants whose scores on parent and teacher-reported normative rating scales exceeded the cutoff and who met the criteria for ADHD using the computerized Kiddie Schedule for Affective Disorders and Schizophrenia for School-Age Children (KSADS) interview. Participants who had met diagnostic criteria in the past but did not at the time of evaluation were also still considered to have ADHD if they remained on medication and demonstrated elevated attentional problems according to the teacher-reported Brief Problem Monitor (BPM) measure.

*ABCD matched case-control subsample.*

In the ABCD cohort, participants identified as ADHD cases (N=540) were matched to two randomly-selected non-ADHD cases from within the same sample. These children were matched exactly on sex, race, ethnicity, and study site and were anti-matched on family (i.e., cases and matched controls were not allowed to share a family ID) using the MatchIt R package (Ho, Imai, King, & Stuart, 2011). A small number of cases who did not achieve a match were then matched without regard to race (N=17) or ethnicity (N=1).

#### Outcomes

*Suicidality.*

In the Oregon ADHD-1000 cohort, measures from the KSADS, the Children's Depression Inventory (CDI), the Structured Clinical Interview for DSM Disorders (SCID), and the Youth Self-Report/Adult Self-Report (YSR/ASR) were used to identify indications of suicidality, including thoughts of death, suicidal ideation, suicide planning, and suicide attempts (current and past). A binary lifetime suicidality variable was created to indicate if a subject had ever experienced any level of suicidality. In the ABCD cohort, items with similar content from the KSADS and the Child Behavior Checklist were used. Again, a binary variable was created to indicate if a subject ever reported having any level of suicidality currently or in the past.

*Alcohol and marijuana use.*

In the Oregon ADHD-1000, alcohol and marijuana use was based on the Customary Drinking and Drug Use Record (CDDR), the Timeline Followback (TLFB), and the Teen Substance Use survey (Nigg et al., 2023).

In the ABCD cohort, marijuana and alcohol use was determined based on positive responses to items that signify use on the following: Youth Mid-Year Phone Interview (su_y_mypi), Youth Substance Use Interview (su_y_sui), Parent KSADS Substance Use Disorder (su_p_ksads_sud), Parent KSADS Summary Scores (mh_p_ksads_ss).

In both the Oregon ADHD-1000 and ABCD cohorts, subjects were asked about current and lifetime substance use with respect to alcohol and marijuana. This was combined into a binarized variable for each substance in which 1 was assigned if a subject had ever endorsed any use of that substance, and 0 if not. For the OR1000 cohort, substance use encompassed ages 13 to 21; in ABCD this encompassed ages 13 to 16.

#### Risk Factors and Covariates

*Lifetime medication status.*

In both cohorts, during study visits caretakers were asked about the children's current and past use of medication for ADHD. A cumulative, binary variable was created to indicate if a child had any history of ADHD medication use.

*Polygenic risk scores.*

ADHD polygenic risk scores, in both cohorts, were calculated with the PRS-CSx tool (Ge et al., 2019; Ruan et al., 2022) using the ADHD-2022 PGC-iPSYCH genome-wide association study (Demontis et al., 2023) as the discovery data set and linkage disequilibrium reference panels from the 1000 Genomes phase 3. For the ABCD cohort, imputed genotypes provided in the ABCD 5.1 data release (http://dx.doi.org/10.15154/z563-zd24) were used. In the Oregon ADHD-1000, genotypes were imputed using the 1000 Genomes phase 3 reference panel as described previously (Nigg et al., 2018). PLINK (Chang et al., 2015) was subsequently used to calculate individual polygenic risk scores as the dot product of the SNP weights (output from PRS-CSx) and genotypes.

#### Complete list of predictors/outcomes.

*Oregon ADHD-1000.*

*Data availability.* Data were generally utilized according to the first available measurement. In general, the following variables correspond to their baseline measurements in the Oregon ADHD-1000, with a few exceptions: In the case of the Alabama parenting variables (Y2_P_C_ALABAMA_INV, Y2_P_C_ALABAMA_PP, Y2_P_C_ALABAMA_PM, Y2_P_C_ALABAMA_ID), the first set of measurements became available in the second year of the study. In the case of Y5_P_PSS_TOTAL, study year 5 was the first instance in which a sufficient number of data points were available.

*Geocoded environment.*

- adi_wsum (Area Deprivation Index, weighted sum)
- r_ED_nat (Child Opportunity Index – education subscale)
- r_HE_na (Child Opportunity Index – health/environment subscale)
- r_SE_nat (Child Opportunity Index – social/economic subscale)
- r_COI_nat (Child Opportunity Index – total)
- NO2_exposure_avg ($NO_2$ annual average exposure)
- PM2.5_exposure_avg (PM2.5 annual average exposure)
- decile (Lead exposure risk decile)

*Family/social environment/genetic.*

- Y1_P_F_FES_CONF_RS (FES – family conflict subscale)
- Y2_P_C_ALABAMA_PP (Alabama – positive parenting subscale)
- Y2_P_C_ALABAMA_INV (Alabama – involvement subscale)

- Y2_P_C_ALABAMA_PM (Alabama – poor monitoring subscale)
- Y2_P_C_ALABAMA_ID (Alabama – inconsistent discipline subscale)
- Y5_P_PSS_TOTAL (Perceived Stress Scale, sum)
- ADHD_PRSCSX (ADHD polygenic risk score)

*Cognitive.*

- ISSRT2 (Stop signal reaction time)
- SPEED (Speed cognitive factor)
- WM (Working memory cognitive factor)
- AROUSAL (Arousal cognitive factor)

*Perinatal/household/family history.*

- birth_weight_oz (Birth weight)
- c_devhx_birth (Premature birth)
- BM_AGE_BIRTH (Biological mother's age at child's birth)
- PRIME_INCOME (Household income)
- Y1_HIGHEST_REL_JOB_SCORE (Highest household job score)
- devhx_edu (Highest household education level)
- Y1_Bm_S_Ksadscid_Adhd (Biological mother's ADHD status, binarized as 'yes' against all other values)

*ABCD 5.1*

*Data availability.* Data were generally utilized according to the first available measurement. In general, the following variables correspond to their baseline measurements in the ABCD 5.1 dataset, with a few exceptions: In the case of three variables (mnbs_ss_mean_all, mnbs_ss_ed_support, and pstr_ss_pr), the first set of measurements were not available until the 3-year followup visit (event name: 3_year_follow_up_y_arm_1).

*Geocoded environment.*

- reshist_addr1_adi_wsum (Area Deprivation Index, weighted sum)
- reshist_addr1_coi_r_ed_nat (Child Opportunity Index – education subscale)
- reshist_addr1_coi_r_he_nat (Child Opportunity Index – health/environment subscale)
- eshist_addr1_coi_r_se_nat (Child Opportunity Index – social/economic subscale)

- reshist_addr1_coi_r_coi_nat (Child Opportunity Index – total)
- reshist_addr1_no2_2016_aavg ($NO_2$ annual average exposure)
- reshist_addr1_pm252016aa (PM2.5 annual average exposure)
- reshist_addr1_leadrisk (Lead exposure risk)

*Family/social environment/genetic*.

- fes_p_ss_fc (FES - family conflict subscale)
- crpbi_y_ss_parent (Child's Report of Parent Behavior Inventory – mean score)
- mnbs_ss_mean_all (Multidimensional Neglectful Behavior Scale, mean)
- mnbs_ss_ed_support (Multidimensional Neglectful Behavior Scale – educational support subscale)
- pmq_y_ss_mean (Parental Monitoring Questionnaire – mean)
- pstr_ss_pr (Perceived Stress Scale, sum)
- ADHDPRS_PRSCSX (ADHD polygenic risk score)

*Cognitive*.

- tfmri_sst_all_beh_total_issrt (Stop signal reaction time)
- nihtbx_flanker_uncorrected (Flanker task)
- nihtbx_list_uncorrected (List sorting task)
- nihtbx_pattern_uncorrected (Pattern comparison processing speed task)

*Perinatal/household/family history*.

- birth_weight (Birth weight)
- gest_age (Gestational age)
- maternal_age_birth (Biological mother's age at child's birth)
- demo_prnt_ed_v2 (Parent education)
- demo_comb_income_v2 (Household income)
- asr_scr_adhd_t (Parent self-reported ADHD symptoms)

#### Missing data.

A breakdown of the available number of observations for all traits, predictors, and outcome variables in each cohort is provided below (**Tables S1 - S4),** with additional outcome reporting statistics in **Table S5.**

| | **Age 9** | **Age 10** | **Age 11** | **Age 12** | **Age 13** | **Age 14-16** |
|---|---|---|---|---|---|---|
| **KSADS Inattention** | 441 | 984 | 1058 | 1090 | 842 | 534 |
| **KSADS Hyperactivity** | 434 | 973 | 1056 | 1090 | 842 | 534 |
| **CBCL Irritability** | 445 | 999 | 1075 | 1104 | 868 | 542 |
| **BPM Attention Mean** | 172 | 446 | 520 | 413 | 213 | 148 |
| **CBCL Inattention** | 445 | 999 | 1075 | 1103 | 868 | 542 |
| **CBCL ADHD** | 445 | 999 | 1075 | 1103 | 868 | 542 |

Table S1. Number of data points available in each age bin for each trait considered in the ABCD cohort out of 1,252 possible subjects.

| | **Age 7/8** | **Age 9** | **Age 10** | **Age 11** | **Age 12** | **Age 13** | **Age 14** | **Age 15** | **Age 16** | **Age 17** | **Age 18-23** |
|---|---|---|---|---|---|---|---|---|---|---|---|
| **ADHDRS Inattention** | 431 | 457 | 479 | 492 | 474 | 409 | 397 | 367 | 286 | 241 | 251 |
| **ADHDRS Hyperactivity** | 431 | 457 | 479 | 492 | 474 | 409 | 397 | 367 | 286 | 241 | 251 |
| **ADHDRS Total Sum** | 431 | 457 | 479 | 492 | 474 | 409 | 397 | 367 | 286 | 241 | 251 |
| **KSADS Irritability** | 438 | 436 | 431 | 383 | 294 | 284 | 244 | 196 | 200 | 157 | 237 |
| **KSADS Inattention** | 438 | 436 | 431 | 383 | 293 | 284 | 244 | 198 | 200 | 158 | 232 |
| **KSADS Hyperactivity** | 438 | 436 | 431 | 383 | 293 | 284 | 244 | 198 | 200 | 157 | 232 |
| **KSADS Total Sum** | 438 | 436 | 431 | 383 | 293 | 284 | 244 | 198 | 200 | 158 | 232 |

| | | | | | | | | | | | |
|---|---|---|---|---|---|---|---|---|---|---|---|
| **TMCQ/EATQ Anger** | 416 | 412 | 408 | 366 | 317 | 326 | 336 | 319 | 271 | 238 | 262 |

Table S2. Number of data points available in each age bin for each trait considered in the Oregon-ADHD-1000 cohort out of 1,215 possible subjects.

| **ABCD - Predictors** | | |
|---|---|---|
| **Variable** | **Type** | **Number (%) missing** |
| ADI WSUM | geocoded | 86 (6.87%) |
| ED NAT | geocoded | 117 (9.35%) |
| HE NAT | geocoded | 117 (9.35%) |
| SE NAT | geocoded | 117 (9.35%) |
| COI NAT | geocoded | 117 (9.35%) |
| NO2 Exp | geocoded | 67 (5.35%) |
| Decile | geocoded | 67 (5.35%) |
| FES | family/social | 0 |
| PSS | family/social | 202 (16.13%) |
| Positive Parenting | family/social | 4 (0.319%) |
| Parental Involvement | family/social | 172 (13.74%) |
| Neglectful Behavior | family/social | 172 (13.74%) |
| Parental Monitoring (PMQ) | family/social | 2 (0.16%) |
| ADHD PRS | family/social | 21 (1.68%) |
| Cognitive - Speed (Pattern) | cognitive | 16 (1.28%) |
| Cognitive - Working Memory | cognitive | 17 (1.36%) |
| Cognitive - Response Inhibition (Flanker) | cognitive | 16 (1.28%) |
| SSRT | cognitive | 40 (3.19%) |
| **ABCD - Outcomes** | | |
| **Variable** | **Type** | **Number (%) missing** |
| Suicidality | outcome | 21 (1.68%) |
| Alcohol use | outcome | 291 (23.24%) |
| Marijuana use | outcome | 257 (20.53%) |
| New disorder onset* | outcome | 124 (9.9%) |

Table S3. Proportion of missing data in the ABCD cohort for predictors and outcomes. Number missing is out of total N=1252.

* numbers of observations were the same for all disorder onset measures.

| **Oregon-ADHD-1000 - Predictors** | | |
|---|---|---|
| **Variable** | **Type** | **Number (%) missing** |
| Env. Exposures and Opportunity* | geocoded | 2 (0.165%) |
| FES | family/social | 410 (33.74%) |

| | | |
|---|---|---|
| PSS | family/social | 886 (72.92%) |
| Alabama Parenting Questionnaire (APQ)** | family/social | 822 (67.65%) |
| ADHD PRS | family/social | 443 (36.46%) |
| Cognitive Speed, WM, Arousal | cognitive | 374 (30.78%) |
| SSRT | cognitive | 449 (36.95%) |
| **Oregon-ADHD-1000 - Outcomes** | | |
| **Variable** | **Type** | **Number (%) missing** |
| Suicidality | outcome | 697 (57.37%) |
| Alcohol use | outcome | 731 (60.16%) |
| Marijuana use | outcome | 730 (60.08%) |
| New disorder onset*** | outcome | 682 (56.13%) |

Table S4. Proportion of missing data in the Oregon-ADHD-1000 cohort for predictors and outcomes. Number missing is out of total N=1215.

* numbers of observations were the same for all geocoded measures.

** numbers of observations were the same for all APQ measures.

*** numbers of observations were the same for all disorder onset measures.

| **ABCD Outcome Reporting** | | | | | | | |
|---|---|---|---|---|---|---|---|
| | **Age 13** | **Age 14** | **Age 15** | **Age 16** | | | **Ever** |
| **Suicidality** | 125 | 91 | 32 | 1 | | | 251 |
| **Marijuana Use** | 12 | 10 | 1 | 0 | | | 21 |
| **Alcohol Use** | 12 | 7 | 2 | 0 | | | 12 |
| **Oregon-ADHD-1000 Outcome Reporting** | | | | | | | |
| | **Age 13** | **Age 14** | **Age 15** | **Age 16** | **Age 17** | **Age 18** | **Ever** |
| **Suicidality** | 80 | 103 | 99 | 82 | 75 | 65 | 208 |
| **Marijuana Use** | 7 | 22 | 37 | 60 | 74 | 69 | 141 |
| **Alcohol Use** | 8 | 25 | 45 | 56 | 84 | 92 | 175 |

Table S5. The number of participants in both cohorts who reported any marijuana use, alcohol use, and any suicidality at the age bins of 13-16 (in the case of ABCD) and age bins of 13-18 (in the case of Oregon-ADHD-1000), and the total number who reported any use across all ages.

#### Additional Symptom Measures.

In the Oregon ADHD-1000 cohort, in addition to the KSADS irritability measure, we examined a revised anger scale from either the Temperament in Middle Childhood Questionnaire or the Early Adolescent Temperament Questionnaire, depending on the child's age at a particular study year (Ellis & Rothbart, 2001; Kozlowski, Antovich, Karalunas, & Nigg, 2022; Kozlowski, Morton, Nigg, & Karalunas, 2025). We also examined the KSADS-measured inattention and hyperactivity results.

The following additional ADHD symptoms measured were examined in the ABCD cohort: the attention problems raw score and the ADHD DSM-oriented raw score from the Child Behavior Checklist (CBCL), as well as the teacher-reported attention problems mean score from the Brief Problem Monitor (BPM).

Results for these additional symptom measures are presented in **Tables S20-S27.**

### Supplemental results.

#### Growth Model Fit Statistics

Fit statistics for all symptom measures used in the primary analyses are shown in **Table S6**. Trajectory plots are shown in **Figures S1 – S3**.

| **Oregon ADHD-1000** | | | | |
|---|---|---|---|---|
| **Trait** | **TLI** | **CFI** | **RMSEA** | **SRMR** |
| ADHDRS – Inattention | 0.990 | 0.987 | 0.021 | 0.036 |
| ADHDRS – Hyperactivity | 0.982 | 0.978 | 0.026 | 0.036 |
| | **BIC** | | **LL** | |
| KSADS - Irritability | 3856.530 | | -1910.509 | |
| **ABCD** | | | | |
| **Trait** | **TLI** | **CFI** | **RMSEA** | **SRMR** |
| KSADS - Inattention | 1.0 | 1.0 | 0.0 | 0.028 |
| KSADS - Hyperactivity | 0.973 | 0.964 | 0.038 | 0.057 |
| CBCL - Irritability | 0.998 | 0.997 | 0.013 | 0.030 |

Table S6. Growth model fit statistics.

#### Growth Model Figures

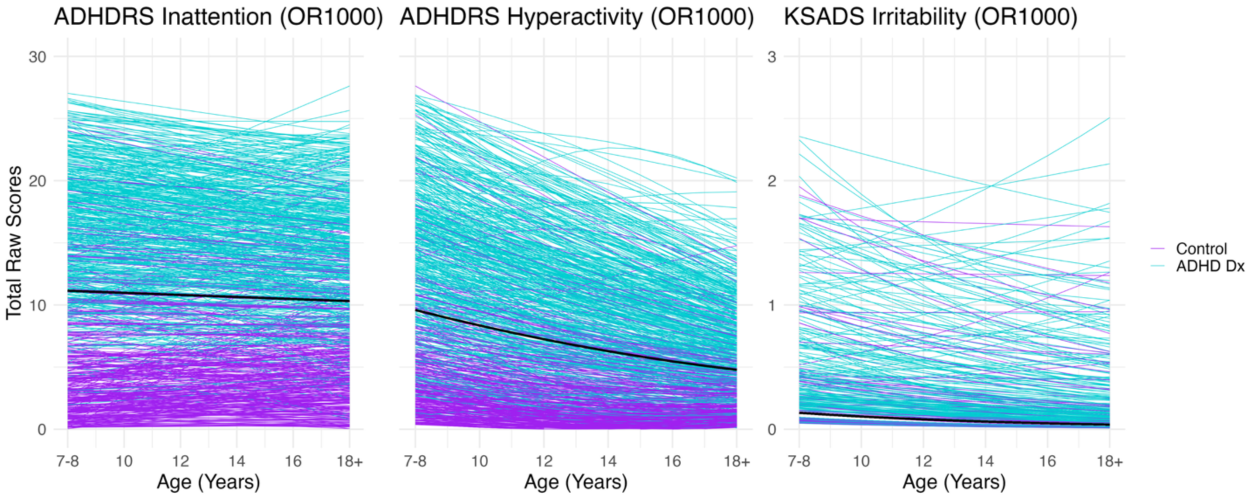

Figure S1. Inattention, hyperactivity, and irritability trajectories in the Oregon ADHD-1000 cohort stratified by case status at baseline.

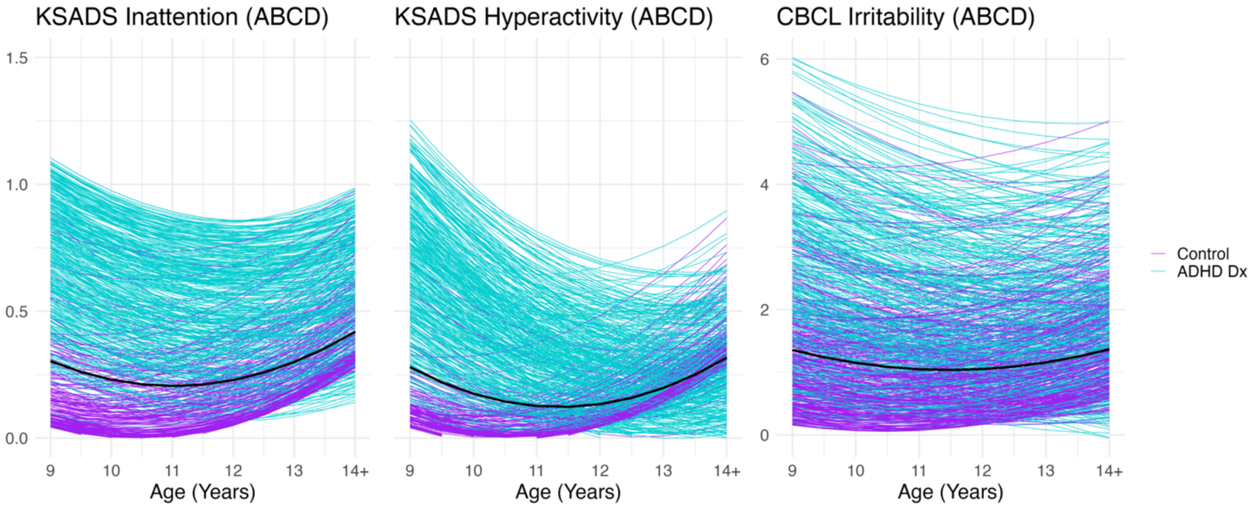

Figure S2. Inattention, hyperactivity, and irritability trajectories in the ABCD cohort stratified by ADHD status at baseline.

#### Trajectory Associations

| | ABCD KSADS Inattention | | OR1000 ADHDRS Inattention | |
|---|---|---|---|---|
| | Intercept | | Intercept | |
| | **Est. (SE)** | **P-value** | **Est. (SE)** | **P-value** |
| ADI | 0.042 (0.027) | 0.117 | 0.048 (0.028) | 0.093 |
| COI | 0.042 (0.028) | 0.141 | -0.006 (0.03) | 0.845 |
| ED | 0.01 (0.029) | 0.724 | -0.05 (0.029) | 0.092 |
| HE | 0.043 (0.028) | 0.12 | 0.046 (0.029) | 0.116 |
| SE | 0.046 (0.029) | 0.106 | 0.006 (0.03) | 0.84 |
| NO2 Exp. | -0.009 (0.029) | 0.763 | 0.015 (0.032) | 0.626 |
| PM2.5 Exp. | -0.031 (0.029) | 0.284 | 0.019 (0.031) | 0.539 |
| Decile | -0.065 (0.028) | 0.02 | -0.054 (0.03) | 0.073 |
| | **ABCD KSADS Hyperactivity** | | **OR1000 ADHDRS Hyperactivity** | |
| | **Intercept** | | **Intercept** | |
| | **Est. (SE)** | **P-value** | **Est. (SE)** | **P-value** |
| ADI | 0.046 (0.027) | 0.095 | 0.081 (0.029) | 0.005 |
| COI | 0.025 (0.028) | 0.373 | -0.052 (0.03) | 0.088 |
| ED | -0.001 (0.029) | 0.981 | -0.103 (0.03) | 0.001 |
| HE | 0.023 (0.029) | 0.418 | 0.027 (0.03) | 0.38 |
| SE | 0.032 (0.029) | 0.275 | -0.031 (0.03) | 0.306 |
| NO2 Exp. | 0.01 (0.028) | 0.727 | 0.02 (0.027) | 0.45 |
| PM2.5 Exp. | -0.023 (0.028) | 0.418 | 0.006 (0.029) | 0.837 |
| Decile | -0.046 (0.029) | 0.115 | -0.031 (0.03) | 0.299 |
| | **ABCD CBCL Irritability** | | **OR1000 KSADS Irritability** | |
| | **Intercept** | | **Intercept** | |
| | **Est. (SE)** | **P-value** | **Est. (SE)** | **P-value** |
| ADI | 0.048 (0.027) | 0.07 | 0.074 (0.027) | 0.006 |
| COI | 0.026 (0.029) | 0.376 | -0.068 (0.03) | 0.026 |
| ED | -0.007 (0.03) | 0.818 | -0.085 (0.03) | 0.005 |
| HE | 0.004 (0.029) | 0.879 | -0.059 (0.03) | 0.047 |
| SE | 0.036 (0.03) | 0.226 | -0.054 (0.029) | 0.067 |
| NO2 Exp. | -0.008 (0.028) | 0.784 | -0.006 (0.037) | 0.879 |
| PM2.5 Exp. | -0.001 (0.031) | 0.966 | 0.028 (0.027) | 0.291 |
| Decile | -0.006 (0.029) | 0.845 | -0.015 (0.028) | 0.583 |

Table S7. Associations between baseline (intercept) ADHD inattention, hyperactivity, and irritability symptoms in the ABCD and OR-1000 cohorts and geocoded environmental and community-level exposures.

| | ABCD KSADS Inattention | | OR1000 ADHDRS Inattention | |
|---|---|---|---|---|
| | Intercept | | Intercept | |
| | **Est. (SE)** | **P-value** | **Est. (SE)** | **P-value** |
| Family Conflict (FES) | 0.261 (0.028) | < 0.001 | 0.172 (0.034) | < 0.001 |
| Perceived Stress (PSS) | 0.205 (0.023) | < 0.001 | 0.245 (0.05) | < 0.001 |
| Positive Parenting* | -0.106 (0.03) | < 0.001 | -0.069 (0.046) | 0.133 |
| Educational support / Involvement* | -0.038 (0.032) | 0.227 | -0.198 (0.044) | < 0.001 |
| Parental Monitoring* | -0.149 (0.03) | < 0.001 | 0.17 (0.048) | < 0.001 |
| OR-ADHD-1000: Inconsistent Discipline* | -- | -- | 0.243 (0.046) | < 0.001 |
| ABCD: Neglectful Behavior* | -0.099 (0.032) | 0.002 | -- | -- |
| ADHD PRS | 0.098 (0.028) | < 0.001 | 0.227 (0.032) | < 0.001 |
| | **ABCD KSADS Hyperactivity** | | **OR1000 ADHDRS Hyperactivity** | |
| | **Intercept** | | **Intercept** | |
| | **Est. (SE)** | **P-value** | **Est. (SE)** | **P-value** |
| Family Conflict | 0.263 (0.029) | < 0.001 | 0.154 (0.034) | < 0.001 |
| Perceived Stress | 0.191 (0.024) | < 0.001 | 0.228 (0.053) | < 0.001 |
| Positive Parenting* | -0.1 (0.031) | 0.001 | -0.015 (0.047) | 0.748 |
| Educational Support / Involvement* | -0.03 (0.03) | 0.324 | -0.165 (0.043) | < 0.001 |
| Parental Monitoring* | -0.136 (0.031) | < 0.001 | 0.079 (0.044) | 0.073 |
| OR-ADHD-1000: Inconsistent Discipline* | -- | -- | 0.189 (0.047) | < 0.001 |
| ABCD: Neglectful Behavior* | -0.088 (0.033) | 0.007 | -- | -- |
| ADHD PRS | 0.103 (0.029) | < 0.001 | 0.259 (0.032) | < 0.001 |
| | **ABCD CBCL Irritability** | | **OR1000 KSADS Irritability** | |
| | **Intercept** | | **Intercept** | |
| | **Est. (SE)** | **P-value** | **Est. (SE)** | **P-value** |
| Family Conflict | 0.356 (0.026) | < 0.001 | 0.216 (0.034) | < 0.001 |
| Perceived Stress | 0.218 (0.025) | < 0.001 | 0.179 (0.052) | 0.001 |
| Positive Parenting* | -0.072 (0.03) | 0.018 | -0.025 (0.051) | 0.616 |
| Educational Support / Involvement* | -0.062 (0.032) | 0.057 | -0.165 (0.044) | < 0.001 |
| Parental Monitoring* | -0.066 (0.029) | 0.022 | 0.124 (0.048) | 0.009 |

| | | | | |
|---|---|---|---|---|
| OR-ADHD-1000: Inconsistent Discipline* | -- | -- | 0.25 (0.042) | < 0.001 |
| ABCD: Neglectful Behavior* | -0.059 (0.033) | 0.069 | -- | -- |
| ADHD PRS | 0.069 (0.029) | 0.016 | 0.158 (0.032) | < 0.001 |

Table S8. Associations between baseline (intercept) ADHD inattention, hyperactivity, and irritability symptoms and predictors related to family and social environment in the ABCD and Oregon ADHD-1000 cohorts. A double-dash (--) indicates variables present in only one cohort. The parenting variables positive parenting, parental involvement, parental monitoring, and inconsistent discipline in the Oregon-ADHD-1000 cohort were measured using the APQ, while in ABCD positive parenting was measured using the CRPBI, parental involvement was measured using the MNBS educational support subscale, parental monitoring was measured using the PMQ, and neglectful behavior was measured using the MNBS total mean score, for which the APQ had no good analogue. In ABCD, there was no analogue for the APQ measure of inconsistent discipline, thus these two variables are considered separately.  In the Oregon ADHD-1000 cohort, the parental monitoring scale is reversed, hence the opposite direction of effect compared to that in ABCD.

| | **ABCD KSADS Inattention** | | **OR1000 ADHDRS Inattention** | |
|---|---|---|---|---|
| | **Intercept** | | **Intercept** | |
| | **Est. (SE)** | **P-value** | **Est. (SE)** | **P-value** |
| OR-ADHD-1000: Speed* | -- | -- | 0.212 (0.03) | < 0.001 |
| ABCD: Speed* | -0.142 (0.029) | < 0.001 | -- | -- |
| Working Memory* | -0.182 (0.027) | < 0.001 | -0.281 (0.029) | < 0.001 |
| OR-ADHD-1000: Arousal* | -- | -- | -0.289 (0.029) | < 0.001 |
| ABCD: Response Inhibition* | -0.128 (0.031) | < 0.001 | -- | -- |
| SSRT | 0.107 (0.029) | < 0.001 | 0.219 (0.03) | < 0.001 |
| | **ABCD KSADS Hyperactivity** | | **OR1000 ADHDRS Hyperactivity** | |
| | **Intercept** | | **Intercept** | |
| | **Est. (SE)** | **P-value** | **Est. (SE)** | **P-value** |
| OR-ADHD-1000: Speed* | -- | -- | 0.146 (0.032) | < 0.001 |
| ABCD: Speed* | -0.101 (0.031) | 0.001 | -- | -- |
| Working Memory* | -0.168 (0.026) | < 0.001 | -0.222 (0.031) | < 0.001 |
| OR-ADHD-1000: Arousal* | -- | -- | -0.241 (0.03) | < 0.001 |
| ABCD: Response Inhibition* | -0.105 (0.034) | 0.002 | -- | -- |
| SSRT | 0.116 (0.031) | < 0.001 | 0.199 (0.033) | < 0.001 |

| | ABCD CBCL Irritability | | OR1000 KSADS Irritability | |
|---|---|---|---|---|
| | Intercept | | Intercept | |
| | Est. (SE) | P-value | Est. (SE) | P-value |
| OR-ADHD-1000: Speed* | -- | -- | 0.057 (0.035) | 0.109 |
| ABCD: Speed* | -0.045 (0.029) | 0.119 | -- | -- |
| Working Memory* | -0.095 (0.029) | 0.001 | -0.069 (0.033) | 0.033 |
| OR-ADHD-1000: Arousal* | -- | -- | -0.044 (0.036) | 0.22 |
| ABCD: Response Inhibition* | -0.069 (0.031) | 0.026 | -- | -- |
| SSRT | 0.106 (0.031) | 0.001 | 0.048 (0.033) | 0.139 |

Table S9. Associations between baseline (intercept) ADHD inattention, hyperactivity, and irritability symptoms in the ABCD and OR-1000 cohorts and cognitive predictors. A double-dash (--) indicates variables present in only one cohort. *In the Oegon-ADHD-1000 cohort, speed, working memory, and cognitive arousal are composite scores made up of scores from several tasks in each domain. In ABCD, speed was measured using the NIH toolbox Pattern Comparison Task, working memory using the NIH Toolbox List Sorting Task, and Response Inhibition using the Flanker task. Only working memory and SSRT are considered harmonious between the cohorts, so speed, arousal, and response inhibition are considered separately.

| | ABCD KSADS Inattention | | | | OR1000 ADHDRS Inattention | | | |
|---|---|---|---|---|---|---|---|---|
| | Intercept | | | | Intercept | | | |
| | Odds Ratio | Lower CI | Upper CI | P-val | Odds Ratio | Lower CI | Upper CI | P-val |
| Alcohol Usage | 0.865 | 0.429 | 1.746 | 0.686 | 0.834 | 0.694 | 1.003 | 0.054 |
| Marijuana Usage | 1.021 | 0.661 | 1.575 | 0.927 | 0.944 | 0.773 | 1.151 | 0.549 |
| Suicidality | 1.078 | 0.943 | 1.231 | 0.272 | 1.147 | 0.968 | 1.361 | 0.100 |
| Mood Disorder Onset | 1.724 | 1.204 | 2.469 | 0.003 | 1.075 | 0.894 | 1.293 | 0.426 |
| Anxiety Disorder Onset | 1.737 | 1.312 | 2.299 | < 0.001 | 1.225 | 1.004 | 1.495 | 0.046 |
| Disruptive Disorder Onset | 2.064 | 1.632 | 2.612 | < 0.001 | 1.261 | 0.977 | 1.629 | 0.072 |

| | | | | | | | | |
|---|---|---|---|---|---|---|---|---|
| Any New Disorder Onset | 1.861 | 1.558 | 2.223 | < 0.001 | 1.164 | 1.572 | 3.944 | 0.082 |
| | **ABCD KSADS Hyperactivity** | | | | **OR1000 ADHDRS Hyperactivity** | | | |
| | **Intercept** | | | | **Intercept** | | | |
| | **Odds Ratio** | **Lower CI** | **Upper CI** | **P-val** | **Odds Ratio** | **Lower CI** | **Upper CI** | **P-val** |
| Alcohol Usage | 0.862 | 0.394 | 1.884 | 0.710 | 0.933 | 0.781 | 1.114 | 0.45 |
| Marijuana Usage | 0.931 | 0.645 | 1.344 | 0.700 | 1.072 | 0.886 | 1.297 | 0.484 |
| Suicidality | 1.081 | 0.944 | 1.238 | 0.261 | 1.149 | 0.976 | 1.353 | 0.107 |
| Mood Disorder Onset | 1.612 | 1.125 | 2.307 | 0.009 | 1.050 | 0.868 | 1.270 | 0.603 |
| Anxiety Disorder Onset | 1.572 | 1.230 | 2.010 | < 0.001 | 1.215 | 0.990 | 1.490 | 0.058 |
| Disruptive Disorder Onset | 1.829 | 1.478 | 2.263 | < 0.001 | 1.087 | 0.851 | 1.389 | 0.485 |
| Any New Disorder Onset | 1.737 | 1.467 | 2.056 | < 0.001 | 1.072 | 0.898 | 1.280 | 0.417 |
| | **ABCD CBCL Irritability** | | | | **OR1000 KSADS Irritability** | | | |
| | **Intercept** | | | | **Intercept** | | | |
| | **Odds Ratio** | **Lower CI** | **Upper CI** | **P-val** | **Odds Ratio** | **Lower CI** | **Upper CI** | **P-val** |
| Alcohol Usage | 0.612 | 0.232 | 1.617 | 0.322 | 1.113 | 0.715 | 1.734 | 0.635 |
| Marijuana Usage | 0.819 | 0.490 | 1.367 | 0.445 | 1.444 | 0.930 | 2.247 | 0.101 |
| Suicidality | 1.066 | 0.931 | 1.221 | 0.355 | 2.312 | 1.444 | 3.718 | < 0.001 |
| Mood Disorder Onset | 2.563 | 1.885 | 3.486 | < 0.001 | 1.531 | 1.012 | 2.305 | 0.04 |
| Anxiety Disorder Onset | 1.694 | 1.334 | 2.149 | < 0.001 | 1.536 | 0.997 | 2.372 | 0.052 |

| | | | | | | | | |
|---|---|---|---|---|---|---|---|---|
| Disruptive Disorder Onset | 1.727 | 1.462 | 2.040 | < 0.001 | 1.821 | 1.040 | 2.617 | 0.034 |
| Any New Disorder Onset | 1.958 | 1.675 | 2.289 | < 0.001 | 0.979 (1.108) | 1.193 | 2.788 | 0.005 |

Table S10. Associations between baseline (intercept) ADHD inattention, hyperactivity, and irritability symptoms in the ABCD and OR-1000 cohorts and behavioral outcomes.

| | **ABCD KSADS Inattention** | | **OR1000 ADHDRS Inattention** | |
|---|---|---|---|---|
| | **Slope** | | **Slope** | |
| | **Est. (SE)** | **P-value** | **Est. (SE)** | **P-value** |
| ADI | -0.015 (0.014) | 0.275 | 0.029 (0.026) | 0.274 |
| COI | 0.058 (0.015) | < 0.001 | -0.048 (0.024) | 0.041 |
| ED | 0.052 (0.016) | < 0.001 | -0.037 (0.023) | 0.105 |
| HE | 0.04 (0.014) | 0.004 | -0.037 (0.024) | 0.134 |
| SE | 0.056 (0.015) | < 0.001 | -0.044 (0.023) | 0.061 |
| NO2 Exp. | 0 (0.014) | 0.984 | -0.006 (0.024) | 0.807 |
| PM2.5 Exp. | 0.01 (0.014) | 0.488 | 0.015 (0.023) | 0.516 |
| Decile | -0.03 (0.014) | 0.034 | 0.047 (0.022) | 0.034 |
| | **ABCD KSADS Hyperactivity** | | **OR1000 ADHDRS Hyperactivity** | |
| | **Slope** | | **Slope** | |
| | **Est. (SE)** | **P-value** | **Est. (SE)** | **P-value** |
| ADI | -0.012 (0.009) | 0.178 | 0.003 (0.025) | 0.917 |
| COI | 0.041 (0.009) | < 0.001 | 0.011 (0.022) | 0.634 |
| ED | 0.036 (0.009) | < 0.001 | 0.014 (0.022) | 0.534 |
| HE | 0.029 (0.008) | 0.001 | -0.008 (0.021) | 0.721 |
| SE | 0.041 (0.009) | < 0.001 | 0.012 (0.022) | 0.587 |
| NO2 Exp. | 0.01 (0.009) | 0.25 | 0.009 (0.022) | 0.68 |
| PM2.5 Exp. | -0.002 (0.009) | 0.787 | 0.019 (0.021) | 0.364 |
| Decile | -0.018 (0.009) | 0.047 | 0.006 (0.021) | 0.762 |
| | **ABCD CBCL Irritability** | | **OR1000 KSADS Irritability** | |
| | **Slope** | | **Slope** | |
| | **Est. (SE)** | **P-value** | **Est. (SE)** | **P-value** |
| ADI | -0.011 (0.019) | 0.562 | 0.021 (0.02) | 0.306 |
| COI | 0.057 (0.019) | 0.002 | -0.013 (0.021) | 0.55 |
| ED | 0.053 (0.02) | 0.008 | -0.015 (0.022) | 0.509 |
| HE | 0.036 (0.018) | 0.05 | -0.016 (0.021) | 0.461 |
| SE | 0.058 (0.019) | 0.003 | -0.009 (0.021) | 0.665 |

| | | | | |
|---|---|---|---|---|
| NO2 Exp. | -0.02 (0.019) | 0.307 | 0.017 (0.021) | 0.418 |
| PM2.5 Exp. | -0.002 (0.019) | 0.91 | 0.03 (0.02) | 0.135 |
| Decile | -0.019 (0.018) | 0.308 | 0.044 (0.022) | 0.05 |

Table S11. Associations between trajectory (slope) ADHD inattention, hyperactivity, and irritability symptoms in the ABCD and OR-1000 cohorts and geocoded environmental and community-level exposures.

| | **ABCD KSADS Inattention** | | **OR1000 ADHDRS Inattention** | |
|---|---|---|---|---|
| | **Slope** | | **Slope** | |
| | **Est. (SE)** | **P-value** | **Est. (SE)** | **P-value** |
| Family Conflict (FES) | -0.014 (0.015) | 0.36 | 0.009 (0.024) | 0.717 |
| Perceived Stress (PSS) | 0.031 (0.015) | 0.035 | 0.015 (0.032) | 0.64 |
| Positive Parenting* | -0.002 (0.015) | 0.876 | 0.025 (0.029) | 0.397 |
| Educational support / Involvement* | -0.015 (0.015) | 0.304 | -0.011 (0.032) | 0.729 |
| Parental Monitoring* | -0.005 (0.015) | 0.754 | -0.024 (0.026) | 0.356 |
| OR-ADHD-1000: Inconsistent Discipline* | -- | -- | 0.028 (0.029) | 0.334 |
| ABCD: Neglectful Behavior* | -0.004 (0.015) | 0.797 | -- | -- |
| ADHD PRS | -0.03 (0.013) | 0.024 | 0.021 (0.025) | 0.406 |
| | **ABCD KSADS Hyperactivity** | | **OR1000 ADHDRS Hyperactivity** | |
| | **Slope** | | **Slope** | |
| | **Est. (SE)** | **P-value** | **Est. (SE)** | **P-value** |
| Family Conflict (FES) | -0.005 (0.011) | 0.638 | -0.032 (0.023) | 0.172 |
| Perceived Stress (PSS) | 0.009 (0.009) | 0.339 | 0.012 (0.033) | 0.717 |
| Positive Parenting | 0.003 (0.01) | 0.784 | -0.024 (0.029) | 0.408 |
| Educational support / Involvement | 0.009 (0.009) | 0.344 | -0.031 (0.028) | 0.277 |
| Parental Monitoring | -0.002 (0.011) | 0.865 | 0.038 (0.025) | 0.133 |

| | | | | |
|---|---|---|---|---|
| OR-ADHD-1000: Inconsistent Discipline | -- | -- | 0.028 (0.029) | 0.334 |
| ABCD: Neglectful Behavior | 0.017 (0.01) | 0.082 | -- | -- |
| ADHD PRS | -0.03 (0.013) | 0.024 | 0.023 (0.024) | 0.329 |
| | **ABCD CBCL Irritability** | | **OR1000 KSADS Irritability** | |
| | **Slope** | | **Slope** | |
| | **Est. (SE)** | **P-value** | **Est. (SE)** | **P-value** |
| Family Conflict (FES) | 0.002 (0.022) | 0.916 | 0.001 (0.024) | 0.969 |
| Perceived Stress (PSS) | 0.025 (0.018) | 0.166 | 0.039 (0.032) | 0.222 |
| Positive Parenting | -0.012 (0.023) | 0.591 | 0.024 (0.022) | 0.27 |
| Educational support / Involvement | -0.054 (0.022) | 0.015 | -0.041 (0.025) | 0.105 |
| Parental Monitoring | 0.009 (0.02) | 0.669 | 0.049 (0.025) | 0.047 |
| OR-ADHD-1000: Inconsistent Discipline | -- | -- | 0.011 (0.024) | 0.645 |
| ABCD: Neglectful Behavior | -0.026 (0.022) | 0.238 | -- | -- |
| ADHD PRS | -0.013 (0.019) | 0.484 | 0.006 (0.023) | 0.806 |

Table S12. Associations between trajectory (slope) ADHD inattention, hyperactivity, and irritability symptoms in the ABCD and OR-1000 cohorts and predictors related to family and social environment. A double-dash (--) indicates variables present in only one cohort.

| | **ABCD KSADS Inattention** | | **OR1000 ADHDRS Inattention** | |
|---|---|---|---|---|
| | **Slope** | | **Slope** | |
| | **Est. (SE)** | **P-value** | **Est. (SE)** | **P-value** |
| Speed (ABCD: Pattern Comparison Task) | -0.001 (0.015) | 0.944 | 0.085 (0.024) | < 0.001 |
| Working Memory (ABCD: List Sorting Task) | 0.026 (0.014) | 0.062 | -0.085 (0.023) | < 0.001 |
| Arousal | -- | -- | -0.044 (0.023) | 0.056 |
| Response Inhibition (Flanker Task) | 0.036 (0.015) | 0.016 | -- | -- |
| SSRT | -0.005 (0.017) | 0.752 | 0.013 (0.027) | 0.62 |

| | ABCD KSADS Hyperactivity | | OR1000 ADHDRS Hyperactivity | |
|---|---|---|---|---|
| | Slope | | Slope | |
| | Est. (SE) | P-value | Est. (SE) | P-value |
| Speed (ABCD: Pattern Comparison Task) | -0.002 (0.01) | 0.808 | 0.07 (0.024) | 0.004 |
| Working Memory (ABCD: List Sorting Task) | 0.027 (0.009) | 0.003 | -0.066 (0.022) | 0.003 |
| Arousal | -- | -- | -0.045 (0.021) | 0.031 |
| Response Inhibition (Flanker Task) | 0.017 (0.009) | 0.061 | -- | -- |
| SSRT | -0.011 (0.013) | 0.431 | 0.034 (0.026) | 0.2 |
| | **ABCD CBCL Irritability** | | **OR1000 KSADS Irritability** | |
| | **Slope** | | **Slope** | |
| | **Est. (SE)** | **P-value** | **Est. (SE)** | **P-value** |
| Speed (ABCD: Pattern Comparison Task) | 0.019 (0.02) | 0.333 | 0.008 (0.025) | 0.764 |
| Working Memory (ABCD: List Sorting Task) | 0.027 (0.018) | 0.14 | -0.009 (0.024) | 0.693 |
| Arousal | -- | -- | -0.008 (0.022) | 0.721 |
| Response Inhibition (Flanker Task) | 0.037 (0.023) | 0.102 | -- | -- |
| SSRT | 0.022 (0.021) | 0.275 | -0.006 (0.035) | 0.875 |

Table S13. Associations between trajectory (slope) ADHD inattention, hyperactivity, and irritability symptoms in the ABCD and OR-1000 cohorts and cognitive predictors. A double-dash (--) indicates variables present in only one cohort.

| | ABCD KSADS Inattention | | OR1000 ADHDRS Inattention | |
|---|---|---|---|---|
| | Intercept | | Intercept | |
| | Est. (SE) | P-value | Est. (SE) | P-value |
| Birth Weight | -0.082 (0.032) | 0.01 | -0.017 (0.036) | 0.636 |
| Gestational Age | -0.066 (0.033) | 0.046 | -0.044 (0.034) | 0.187 |
| Biological Mother's Age At Child Birth | -0.113 (0.029) | < 0.001 | -0.151 (0.030) | < 0.001 |
| Highest Household Income | -0.061 (0.029) | 0.033 | -0.097 (0.030) | 0.001 |
| Highest Household Employment | -- | -- | -0.096 (0.029) | 0.001 |
| Highest Household Education | 0.007 (0.026) | 0.781 | -0.122 (0.034) | < 0.001 |

| | | | | |
|---|---|---|---|---|
| Biological Mother ADHD Diagnosis | -- | -- | 0.173 (0.031) | < 0.001 |
| Parental ADHD Symptoms | 0.402 (0.026) | < 0.001 | -- | -- |
| | **ABCD KSADS Hyperactivity** | | **OR1000 ADHDRS Hyperactivity** | |
| | **Intercept** | | **Intercept** | |
| | **Est. (SE)** | **P-value** | **Est. (SE)** | **P-value** |
| Birth Weight | -0.100 (0.033) | 0.002 | -0.031 (0.037) | 0.394 |
| Gestational Age | -0.064 (0.035) | 0.066 | -0.055 (0.032) | 0.086 |
| Biological Mother's Age At Child Birth | -0.104 (0.031) | 0.001 | -0.152 (0.030) | < 0.001 |
| Highest Household Income | -0.106 (0.031) | 0.001 | -0.137 (0.031) | < 0.001 |
| Highest Household Employment | -- | -- | -0.158 (0.030) | < 0.001 |
| Highest Household Education | -0.029 (0.028) | 0.297 | -0.172 (0.033) | < 0.001 |
| Biological Mother ADHD Diagnosis | -- | -- | 0.154 (0.035) | < 0.001 |
| Parental ADHD Symptoms | 0.378 (0.027) | < 0.001 | -- | -- |
| | **ABCD CBCL Irritability** | | **OR1000 KSADS Irritability** | |
| | **Intercept** | | **Intercept** | |
| | **Est. (SE)** | **P-value** | **Est. (SE)** | **P-value** |
| Birth Weight | -0.021 (0.030) | 0.490 | 0.013 (0.031) | 0.683 |
| Gestational Age | -0.010 (0.029) | 0.720 | -0.005 (0.033) | 0.876 |
| Biological Mother's Age At Child Birth | -0.104 (0.030) | 0.001 | -0.093 (0.034) | 0.006 |
| Highest Household Income | -0.107 (0.032) | 0.001 | -0.142 (0.030) | < 0.001 |
| Highest Household Employment | -- | -- | -0.189 (0.036) | < 0.001 |
| Highest Household Education | -0.031 (0.028) | 0.273 | -0.083 (0.032) | 0.010 |
| Biological Mother ADHD Diagnosis | -- | -- | 0.101 (0.042) | 0.017 |
| Parental ADHD Symptoms | 0.422 (0.028) | < 0.001 | -- | -- |

Table S14. Associations between baseline symptoms (intercept) and potentially confounding risk factors in the ABCD and Oregon ADHD-1000 cohorts. A double-dash (--) indicates variables present in only one cohort.

| | ABCD KSADS Inattention | | OR1000 ADHDRS Inattention | |
|---|---|---|---|---|
| | Slope | | Slope | |
| | Est. (SE) | P-value | Est. (SE) | P-value |
| Birth Weight | 0.019 (0.014) | 0.190 | -0.019 (0.024) | 0.423 |
| Gestational Age | 0.006 (0.014) | 0.656 | -0.004 (0.022) | 0.852 |
| Biological Mother's Age At Child Birth | 0.025 (0.015) | 0.097 | -0.007 (0.027) | 0.792 |
| Highest Household Income | 0.048 (0.015) | 0.001 | -0.053 (0.025) | 0.038 |
| Highest Household Employment | -- | -- | -0.064 (0.029) | 0.025 |
| Highest Household Education | 0.050 (0.014) | < 0.001 | -0.051 (0.023) | 0.028 |
| Biological Mother ADHD Diagnosis | -- | -- | 0.005 (0.032) | 0.875 |
| Parental ADHD Symptoms | -0.004 (0.021) | 0.847 | -- | -- |
| | **ABCD KSADS Hyperactivity** | | **OR1000 ADHDRS Hyperactivity** | |
| | **Slope** | | **Slope** | |
| | **Est. (SE)** | **P-value** | **Est. (SE)** | **P-value** |
| Birth Weight | -0.100 (0.033) | 0.002 | -0.013 (0.024) | 0.604 |
| Gestational Age | -0.001 (0.011) | 0.907 | 0.017 (0.021) | 0.414 |
| Biological Mother's Age At Child Birth | 0.031 (0.010) | 0.001 | 0.001 (0.026) | 0.983 |
| Highest Household Income | 0.038 (0.010) | < 0.001 | -0.035 (0.027) | 0.194 |
| Highest Household Employment | -- | -- | -0.036 (0.033) | 0.272 |
| Highest Household Education | 0.039 (0.009) | < 0.001 | -0.016 (0.023) | 0.474 |
| Biological Mother ADHD Diagnosis | -- | -- | 0.011 (0.042) | 0.79 |
| Parental ADHD Symptoms | -0.003 (0.015) | 0.854 | -- | -- |
| | **ABCD CBCL Irritability** | | **OR1000 KSADS Irritability** | |
| | **Slope** | | **Slope** | |
| | **Est. (SE)** | **P-value** | **Est. (SE)** | **P-value** |
| Birth Weight | 0.017 (0.019) | 0.374 | -0.017 (0.022) | 0.450 |
| Gestational Age | 0.034 (0.019) | 0.078 | -0.005 (0.025) | 0.826 |
| Biological Mother's Age At Child Birth | 0.051 (0.020) | 0.011 | -0.026 (0.024) | 0.285 |

| | | | | |
|---|---|---|---|---|
| Highest Household Income | 0.037 (0.020) | 0.057 | -0.020 (0.025) | 0.408 |
| Highest Household Employment | -- | -- | 0.005 (0.025) | 0.832 |
| Highest Household Education | 0.053 (0.017) | 0.002 | -0.051 (0.025) | 0.037 |
| Biological Mother ADHD Diagnosis | -- | -- | 0.018 (0.023) | 0.437 |
| Parental ADHD Symptoms | -0.006 (0.027) | 0.815 | -- | -- |

Table S15. Associations between symptom trajectories (slope) and potentially confounding risk factors in the ABCD and Oregon ADHD-1000 cohorts. A double-dash (--) indicates variables present in only one cohort.

| | **ABCD KSADS Inattention** | | **OR1000 ADHDRS Inattention** | |
|---|---|---|---|---|
| | **Intercept** | | **Intercept** | |
| | **Est. (SE)** | **P-value** | **Est. (SE)** | **P-value** |
| Family Conflict (FES) | 0.141 (0.028) | < 0.001 | 0.151 (0.032) | < 0.001 |
| Perceived Stress (PSS) | 0.136 (0.024) | < 0.001 | 0.195 (0.053) | < 0.001 |
| Parental Monitoring | -0.106 (0.030) | < 0.001 | 0.140 (0.044) | 0.002 |
| Speed (OR1000 composite) | -- | -- | 0.2p3 (0.030) | < 0.001 |
| Speed (ABCD Pattern Comparison Task) | -0.104 (0.027) | < 0.001 | -- | -- |
| Working Memory (ABCD: List Sorting Task) | -0.167 (0.025) | < 0.001 | -0.267 (0.030) | < 0.001 |
| SSRT | 0.078 (0.027) | 0.004 | 0.207 (0.029) | < 0.001 |
| ADHD PRS | 0.085 (0.030) | 0.004 | 0.197 (0.033) | < 0.001 |
| | **ABCD KSADS Hyperactivity** | | **OR1000 ADHDRS Hyperactivity** | |
| | **Intercept** | | **Intercept** | |
| | **Est. (SE)** | **P-value** | **Est. (SE)** | **P-value** |
| Family Conflict (FES) | 0.145 (0.029) | < 0.001 | 0.127 (0.033) | < 0.001 |

| | | | | | | | | |
|---|---|---|---|---|---|---|---|---|
| Perceived Stress (PSS) | 0.132 (0.025) | | | < 0.001 | 0.171 (0.056) | | | 0.002 |
| Speed (OR1000 composite) | -- | | | -- | 0.129 (0.031) | | | < 0.001 |
| Speed (ABCD Pattern Comparison Task) | 0.055 (0.029) | | | 0.057 | -- | | | -- |
| Response Inhibition (Flanker Task) | -0.087 (0.034) | | | 0.010 | -- | | | -- |
| Working Memory (ABCD: List Sorting Task) | -0.184 (0.026) | | | < 0.001 | -0.198 (0.031) | | | < 0.001 |
| SSRT | 0.111 (0.026) | | | < 0.001 | 0.185 (0.032) | | | < 0.001 |
| ADHD PRS | 0.071 (0.031) | | | 0.023 | 0.219 (0.032) | | | < 0.001 |
| | **ABCD CBCL Irritability** | | | | **OR1000 KSADS Irritability** | | | |
| | **Intercept** | | | | **Intercept** | | | |
| | **Est. (SE)** | | | **P-value** | **Est. (SE)** | | | **P-value** |
| Family Conflict (FES) | 0.262 (0.027) | | | < 0.001 | 0.196 (0.033) | | | < 0.001 |
| Perceived Stress (PSS) | 0.173 (0.027) | | | < 0.001 | 0.144 (0.057) | | | 0.011 |
| | **Odds Ratio** | **Lower CI** | **Upper CI** | **P-value** | **Odds Ratio** | **Lower CI** | **Upper CI** | **P-value** |
| Any New Disorder Onset | 1.361 | 1.493 | 2.391 | < 0.001 | 1.951 | 1.237 | 3.090 | 0.003 |

Table S16. Results of covariate-adjusted analysis for replicated associations with baseline symptoms. Only nominally significant effects found in the primary analyses were subject to covariate-adjusted analysis as follow-up. A double-dash (--) indicates variables present in only one cohort.

| | **ABCD KSADS Inattention** | | **OR1000 ADHDRS Inattention** | |
|---|---|---|---|---|
| | **Slope** | | **Slope** | |
| | **Est. (SE)** | **P-value** | **Est. (SE)** | **P-value** |
| COI | 0.039 (0.019) | 0.04 | -0.032 (0.027) | 0.237 |
| ED | 0.033 (0.018) | 0.072 | *N/A* | *N/A* |
| HE | 0.019 (0.016) | 0.231 | *N/A* | *N/A* |
| SE | 0.035 (0.018) | 0.052 | *N/A* | *N/A* |

| | | | | | | | | |
|---|---|---|---|---|---|---|---|---|
| Decile | -0.071 (0.03) | | | 0.019 | 0.043 (0.023) | | | 0.058 |
| Perceived Stress (PSS) | 0.019 (0.016) | | | 0.233 | *N/A* | | | *N/A* |
| Speed (OR1000 Composite) | -- | | | -- | 0.08 (0.024) | | | 0.001 |
| Working Memory (ABCD: List Sorting Task) | *N/A* | | | *N/A* | -0.078 (0.024) | | | 0.001 |
| Response Inhibition (Flanker Task) | 0.026 (0.016) | | | 0.088 | -- | | | -- |
| LT Medication Status | *N/A* | | | *N/A* | 0.212 (0.034) | | | < 0.001 |
| ADHD PRS | -0.012 (0.015) | | | 0.415 | *N/A* | | | *N/A* |
| | **Odds Ratio** | **Lower CI** | **Upper CI** | **P-value** | **Odds Ratio** | **Lower CI** | **Upper CI** | **P-value** |
| LT Suicidality | *N/A* | | | | 1.223 | 1.046 | 1.431 | 0.012 |
| Mood Disorder Onset | 6.349 | 2.437 | 16.539 | < 0.001 | *N/A* | | | |
| Disruptive Disorder Onset | 3.060 | 1.813 | 5.163 | < 0.001 | 1.557 | 1.146 | 2.117 | 0.005 |
| Any New Disorder Onset | 2.477 | 1.583 | 3.876 | < 0.001 | *N/A* | | | |
| | **ABCD KSADS Hyperactivity** | | | | **OR1000 ADHDRS Hyperactivity** | | | |
| | **Est. (SE)** | | | **P-value** | **Est.** | | | **P-value** |
| COI | 0.023 (0.011) | | | 0.049 | *N/A* | | | *N/A* |
| ED | 0.016 (0.011) | | | 0.135 | *N/A* | | | *N/A* |
| HE | 0.01 (0.009) | | | 0.289 | *N/A* | | | *N/A* |
| SE | 0.024 (0.011) | | | 0.031 | *N/A* | | | *N/A* |
| Decile | -0.004 (0.01) | | | 0.689 | *N/A* | | | *N/A* |
| Speed (OR1000 Composite) | -- | | | -- | 0.067 (0.024) | | | 0.005 |
| Working Memory (ABCD: List Sorting Task) | 0.014 (0.009) | | | 0.145 | -0.065 (0.022) | | | 0.004 |
| Cog, Arousal (OR1000 Composite) | -- | | | -- | -0.043 (0.021) | | | 0.038 |
| LT Medication Status | *N/A* | | | *N/A* | 0.147 (0.029) | | | < 0.001 |
| ADHD PRS | -0.005 (0.009) | | | 0.566 | *N/A* | | | *N/A* |

| | Odds Ratio | Lower CI | Upper CI | P-value | Odds Ratio | Lower CI | Upper CI | P-value |
|---|---|---|---|---|---|---|---|---|
| Alcohol Use | *N/A* | | | | 0.891 | 0.682 | 0.976 | 0.026 |
| Mood Disorder Onset | 7.010 | 1.370 | 35.866 | 0.019 | *N/A* | | | |
| Disruptive Disorder Onset | 4.732 | 2.127 | 10.526 | **< 0.001** | *N/A* | | | |
| Any New Disorder Onset | 3.430 | 1.802 | 6.527 | < 0.001 | *N/A* | | | |
| | **ABCD CBCL Irritability** | | | | **OR1000 KSADS Irritability** | | | |
| | **Est. (SE)** | | | **P-value** | **Est.** | | | **P-value** |
| COI | 0.037 (0.025) | | | 0.135 | *N/A* | | | *N/A* |
| ED | 0.029 (0.025) | | | 0.231 | *N/A* | | | *N/A* |
| HE | 0.014 (0.021) | | | 0.49 | *N/A* | | | *N/A* |
| SE | 0.038 (0.024) | | | 0.115 | *N/A* | | | *N/A* |
| Educational support / Involvement | -0.057 (0.022) | | | 0.01 | *N/A* | | | *N/A* |
| Parental Monitoring | *N/A* | | | *N/A* | 0.055 (0.024) | | | 0.023 |
| LT Medication Status | *N/A* | | | *N/A* | 0.071 (0.031) | | | 0.022 |
| Sex | *N/A* | | | *N/A* | 0.057 (0.024) | | | 0.019 |
| | **Odds Ratio** | **Lower CI** | **Upper CI** | **P-value** | **Odds Ratio** | **Lower CI** | **Upper CI** | **P-value** |
| Mood Disorder Onset | 3.311 | 2.169 | 5.057 | < 0.001 | *N/A* | | | |
| Disruptive Disorder Onset | 2.042 | 1.478 | 2.822 | < 0.001 | 1.717 | 1.256 | 2.350 | 0.001 |
| Any New Disorder Onset | 1.804 | 1.361 | 2.391 | < 0.001 | *N/A* | | | |

Table S17. Results of covariate-adjusted analysis for associations with symptom trajectories (slopes). Only nominally significant effects found in the primary analyses with respect to at least one cohort were subject to covariate-adjusted analysis as follow-up; rows for which a variable existed but was not subject to covariate-adjusted analysis are indicated by italicized N/A. A double-dash (--) indicates variables present in only one cohort.

| | ABCD KSADS Inattention | OR1000 ADHDRS Inattention |
|---|---|---|

| | Intercept | | Intercept | |
|---|---|---|---|---|
| | Est. (SE) | P-value | Est. (SE) | P-value |
| LT Medication Status | 0.189 (0.03) | < 0.001 | 0.618 (0.022) | < 0.001 |
| Sex | 0.035 (0.028) | 0.212 | 0.15 (0.028) | < 0.001 |
| | **ABCD KSADS Hyperactivity** | | **OR1000 ADHDRS Hyperactivity** | |
| | **Intercept** | | **Intercept** | |
| | **Est. (SE)** | **P-value** | **Est. (SE)** | **P-value** |
| LT Medication Status | 0.195 (0.032) | < 0.001 | 0.556 (0.024) | < 0.001 |
| Sex | 0.07 (0.027) | 0.01 | 0.16 (0.028) | < 0.001 |
| | **ABCD CBCL Irritability** | | **OR1000 KSADS Irritability** | |
| | **Intercept** | | **Intercept** | |
| | **Est. (SE)** | **P-value** | **Est. (SE)** | **P-value** |
| LT Medication Status | 0.116 (0.031) | < 0.001 | 0.219 (0.032) | < 0.001 |
| Sex | 0 (0.029) | 0.992 | 0.107 (0.025) | < 0.001 |

Table S18. Associations between baseline (intercept) ADHD inattention, hyperactivity, and irritability symptoms in the ABCD and OR-1000 cohorts and miscellaneous predictors (lifetime medication status and sex).

| | ABCD KSADS Inattention | | OR1000 ADHDRS Inattention | |
|---|---|---|---|---|
| | **Slope** | | **Slope** | |
| | **Est. (SE)** | **P-value** | **Est. (SE)** | **P-value** |
| LT Medication Status | -0.014 (0.015) | 0.372 | 0.211 (0.034) | < 0.001 |
| Sex | 0.023 (0.014) | 0.1 | 0.013 (0.023) | 0.565 |
| | **ABCD KSADS Hyperactivity** | | **OR1000 ADHDRS Hyperactivity** | |
| | **Slope** | | **Slope** | |
| | **Est. (SE)** | **P-value** | **Est. (SE)** | **P-value** |
| LT Medication Status | 0.011 (0.011) | 0.328 | 0.149 (0.029) | < 0.001 |
| Sex | 0.015 (0.009) | 0.089 | 0.034 (0.022) | 0.114 |
| | **ABCD CBCL Irritability** | | **OR1000 KSADS Irritability** | |
| | **Slope** | | **Slope** | |

| | Est. (SE) | P-value | Est. (SE) | P-value |
|---|---|---|---|---|
| LT Medication Status | -0.008 (0.022) | 0.718 | 0.053 (0.023) | 0.019 |
| Sex | -0.029 (0.02) | 0.139 | -0.002 (0.02) | 0.927 |

Table S19. Associations between trajectory (slope) ADHD inattention, hyperactivity, and irritability symptoms in the ABCD and OR-1000 cohorts and miscellaneous predictors (lifetime medication status and sex).

#### Additional Symptom Measure Analyses

Result for additional symptom measures are reported below. Fit statistics and mean growth parameters are shown in **Table S20 and Table S21.**

| Oregon ADHD-1000 | | | | |
|---|---|---|---|---|
| **Trait** | **TLI** | **CFI** | **RMSEA** | **SRMR** |
| KSADS – Inattention | 0.976 | 0.969 | 0.022 | 0.057 |
| KSADS – Hyperactivity | 0.979 | 0.974 | 0.016 | 0.070 |
| TMCQ/EATQ - Anger | 0.982 | 0.976 | 0.024 | 0.074 |
| **ABCD** | | | | |
| **Trait** | **TLI** | **CFI** | **RMSEA** | **SRMR** |
| BPM – Inattention Mean (teacher) | 1.0 | 1.0 | 0.0 | 0.079 |
| CBCL – Inattention problems raw score | 0.994 | 0.991 | 0.029 | 0.025 |
| CBCL – DSM-oriented ADHD raw score | 0.996 | 0.995 | 0.024 | 0.026 |

Table S20. Growth model fit statistics for additional symptom measures.

| Oregon ADHD-1000 | | | | |
|---|---|---|---|---|
| **Symptom** | **Term** | **Est.** | **SE** | **P-value** |
| KSADS - Inattention | I | 0.438 | 0.012 | < 0.001 |
| | S | -0.012 | 0.002 | < 0.001 |
| KSADS - Hyperactivity | I | 0.346 | 0.012 | < 0.001 |
| | S | -0.038 | 0.004 | < 0.001 |
| | Q | 0.001 | 0.00 | < 0.001 |
| KSADS - Total | I | 0.767 | 0.020 | < 0.001 |
| | S | -0.037 | 0.003 | < 0.001 |
| TMCQ/EATQ - Anger raw scores | I | 2.352 | 0.021 | < 0.001 |
| | S | -0.041 | 0.004 | < 0.001 |
| **ABCD** | | | | |
| **Symptom** | | **Est.** | **SE** | **P-value** |
| BPM - Inattention Mean | I | 0.678 | 0.030 | < 0.001 |
| | S | -0.043 | 0.009 | < 0.001 |
| CBCL – Inattention RS | I | 5.288 | 0.174 | < 0.001 |
| | S | -0.487 | 0.080 | < 0.001 |
| | Q | 0.047 | 0.014 | 0.001 |
| CBCL – ADHD RS | I | 4.582 | 0.146 | < 0.001 |
| | S | -0.438 | 0.065 | < 0.001 |
| | Q | 0.034 | 0.012 | 0.003 |

Table S21. Mean growth parameters for additional symptom measures.

#### Oregon ADHD-1000

##### *Oregon-ADHD-1000 TMCQ/EATQ Anger.*

The results of the TMCQ/EATQ anger measure regression analyses were in agreement with those of irritability (both Oregon-ADHD-1000 KSADS and ABCD CBCL). Fewer significant effects were observed with respect to both baseline and over time with the exception of family environment predictors, which were statistically significant over time as well as at baseline; this is notable as this was not generally the case for any other trait examined. The significant effects that were observed were consistent in direction and magnitude with the other related traits.

###### *Community Environment:*

*Baseline.* Following multiple testing corrections, no environmental predictor achieved statistical significance in relation to anger at baseline, although a nominal effect was observed for ED (coeff: -0.08 (0.03), $p = 0.007$; **Table S22).**

*Trajectory.* Following multiple testing corrections, no environmental predictor achieved statistical significance in relation to change in anger measurement over time, although a nominal effect was observed for PM2.5 exposure (coeff: 0.072 (0.025), $p = 0.004$; **Table S22).**

###### *Family Environment:*

*Baseline.* All family and social predictors were significantly associated with TMCQ/EATQ-measured anger at baseline ($p < 0.001$; **Table S22).**

*Trajectory.* All family and social predictors were significantly associated with anger measured over time ($p < 0.001$) except for PSS, positive parenting, and parental monitoring, although PSS showed a nominal effect in the opposite of the expected direction (**Table S22**).

###### *ADHD Polygenic Risk, Medication Status, and Sex:*

*Baseline.* Lifetime medication status (coeff = 0.293 (0.032), $p < 0.001$) and polygenic risk (coeff = 0.208 (0.034), $p < 0.001$) were significantly associated with TMCQ/EATQ-measured anger at baseline (**Table S22**).

*Trajectory.* Only lifetime medication status was significantly associated with change in anger over time (coeff = 0.144 (0.028), $p < 0.001$; **Table S22).**

*Cognition:*

*Baseline.* Following multiple testing corrections, no cognitive predictor achieved statistical significance in relation to anger measured at baseline. Nominal effects were observed for working memory (coeff: -0.099 (0.036), p = 0.006) and cognitive arousal (coeff = -0.103 (0.035), p = 0.003; **Table S22).**

*Trajectory.* Following multiple testing, a significant association was observed between working memory and change in anger over time (coeff = -0.077 (0.025), p = 0.002). A nominal effect was observed for cognitive arousal (coeff: -0.067 (0.025), p = 0.008; **Table S22).**

*Outcomes:*

*Baseline*. Following multiple testing corrections, only suicidality was significantly associated with TMCQ/EATQ-measured anger at baseline (OR = 1.338 [1.118 – 1.600], p = 0.002; **Table S22**).

*Trajectory.* Following multiple testing corrections, no outcomes were observed to be significantly associated with change in anger over time; however, nominal effects were observed for substance use (both alcohol and marijuana) and suicidality (**Table S22**).

| | **OR1000 TMCQ/EATQ Anger** | | | |
|---|---|---|---|---|
| | **Intercept** | | **Slope** | |
| **Community Environment** | **Est. (SE)** | **P-value** | **Est. (SE)** | **P-value** |
| ADI | 0.061 (0.03) | 0.043 | 0.004 (0.025) | 0.868 |
| ED | -0.08 (0.03) | 0.007 | 0.008 (0.026) | 0.762 |
| HE | -0.03 (0.032) | 0.36 | 0.011 (0.030) | 0.706 |
| SE | -0.065 (0.031) | 0.034 | -0.010 (0.027) | 0.716 |
| COI | -0.073 (0.031) | 0.016 | -0.005 (0.027) | 0.853 |
| NO2 | -0.047 (0.027) | 0.08 | -0.002 (0.024) | 0.924 |
| PM2.5 | 0.021 (0.03) | 0.488 | 0.072 (0.025) | 0.004 |
| Decile | 0.001 (0.031) | 0.986 | 0.034 (0.025) | 0.165 |
| **Family Environment** | | | | |
| Family Conflict (FES) | 0.181 (0.036) | < 0.001 | 0.087 (0.025) | 0.001 |
| Perceived Stress (PSS) | 0.192 (0.051) | < 0.001 | 0.104 (0.035) | 0.003 |
| AL Positive Parenting | -0.082(0.049) | 0.093 | 0.040 (0.036) | 0.271 |

| | | | | | | | | |
|---|---|---|---|---|---|---|---|---|
| AL Parental Involvement | -0.159 (0.045) | | | < 0.001 | -0.110 (0.029) | | | < 0.001 |
| AL Poor Monitoring | 0.173 (0.047) | | | < 0.001 | 0.057 (0.027) | | | 0.034 |
| AL Inconsistent Discipline | 0.279 (0.048) | | | < 0.001 | 0.129 (0.032) | | | < 0.001 |
| **ADHD Polygenic Risk, Medication Status, and Sex** | **Est. (SE)** | | | **P-value** | **Est. (SE)** | | | **P-value** |
| ADHD PRS | 0.208 (0.034) | | | < 0.001 | 0.054 (0.028) | | | 0.050 |
| LT Med. Status | 0.293 (0.032) | | | < 0.001 | 0.144 (0.028) | | | < 0.001 |
| Sex | 0.035 (0.029) | | | 0.218 | -0.014 (0.025) | | | 0.577 |
| **Cognition** | **Est. (SE)** | | | **P-value** | **Est. (SE)** | | | **P-value** |
| Cog. Speed (Composite) | 0.057 (0.035) | | | 0.109 | 0.054 (0.025) | | | 0.028 |
| Working Memory (Composite) | -0.099 (0.036) | | | 0.006 | -0.077 (0.025) | | | 0.002 |
| Cog. Arousal (Composite) | -0.103 (0.035) | | | 0.003 | -0.067 (0.025) | | | 0.008 |
| SSRT | 0.095 (0.037) | | | 0.01 | 0.018 (0.029) | | | 0.532 |
| **Outcomes** | **Odds Ratio** | **Lower CI** | **Upper CI** | **P-value** | **Odds Ratio** | **Lower CI** | **Upper CI** | **P-value** |
| Alcohol Use | 0.903 | 0.746 | 1.092 | 0.289 | 1.215 | 1.050 | 1.406 | 0.009 |
| Marijuana Use | 1.053 | 0.863 | 1.285 | 0.610 | 1.220 | 1.051 | 1.416 | 0.009 |
| Suicidality | 1.338 | 1.118 | 1.600 | 0.002 | 1.227 | 1.067 | 1.411 | 0.004 |
| Mood Disorder Onset | 1.133 | 0.929 | 1.383 | 0.217 | 1.113 | 0.930 | 1.332 | 0.242 |
| Anxiety Disorder Onset | 1.126 | 0.913 | 1.389 | 0.264 | 1.232 | 1.020 | 1.489 | 0.030 |
| Disruptive Disorder Onset | 1.394 | 1.080 | 1.799 | 0.011 | 1.268 | 0.938 | 1.714 | 0.123 |
| Any New Disorder Onset | 1.200 | 1.002 | 1.437 | 0.048 | 1.164 | 0.984 | 1.377 | 0.075 |

Table S22. Associations between anger symptoms at baseline (left-most columns) and over time (right-most columns) in the Oregon-ADHD-1000 cohort, and predictor/outcome variables of interest.

##### *Oregon-ADHD-1000 KSADS Inattention.*

The results of the regression analyses conducted with KSADS-measured inattention in the Oregon-ADHD-1000 cohort and our predictors and outcomes of interest were in remarkable agreement with those conducted with ADHD RS-measured inattention, as well as with ABCD KSADS. Family/social environment and cognitive variables stood out as significant associations with baseline symptoms, while very few variables were related to symptom change over time, consistent with other measures of inattention.

*Community Environment:*

Following multiple testing corrections, no environmental predictor achieved statistical significance in relation to KSADS-measured in attention either at baseline or over time.

*Family Environment:*

*Baseline.* All family and social predictors were significantly associated with KSADS - measured inattention at baseline ($p < 0.001$; **Table S23**) except for positive parenting.

*Trajectory.* No family and social predictors were significantly associated with KSADS - measured inattention over time.

*ADHD Polygenic Risk, Medication Status, and Sex:*

*Baseline.* Lifetime medication status (coeff = 0.642 (0.022), $p < 0.001$), polygenic risk (coeff = 0.230 (0.032), $p < 0.001$), and male sex (coeff = 0.162 (0.028), $p < 0.001$) were significantly associated with KSADS-measured inattention at baseline; see **Table S23**.

*Trajectory.* Only lifetime medication status was significantly associated with change in KSADS-measured inattention over time (coeff = 0.174 (0.037), $p < 0.001$; **Table S23)**.

*Cognition:*

*Baseline.* All cognitive predictors were significantly associated with KSADS -measured inattention at baseline ($p < 0.001$; **Table S23**).

*Trajectory.* No cognitive predictors were significantly associated with KSADS -measured inattention over time.

*Outcomes:*

Following multiple testing corrections, no outcomes were significantly associated with KSADS -measured inattention at baseline or over time.

| | **OR1000 KSADS Inattention** | |
|---|---|---|
| | **Intercept** | **Slope** |

| Community Environment | Est. (SE) | | | P-value | Est. (SE) | | | P-value |
|---|---|---|---|---|---|---|---|---|
| ADI | 0.053 (0.028) | | | 0.060 | -0.009 (0.026) | | | 0.722 |
| ED | -0.043 (0.029) | | | 0.141 | -0.013 (0.021) | | | 0.541 |
| HE | 0.037 (0.029) | | | 0.207 | -0.025 (0.022) | | | 0.264 |
| SE | -0.006 (0.029) | | | 0.837 | -0.026 (0.021) | | | 0.226 |
| COI | -0.013 (0.029) | | | 0.652 | -0.027 (0.021) | | | 0.201 |
| NO2 | -0.015 (0.029) | | | 0.622 | 0.030 (0.022) | | | 0.178 |
| PM2.5 | 0.023 (0.029) | | | 0.423 | -0.009 (0.021) | | | 0.675 |
| Decile | -0.063 (0.028) | | | 0.026 | 0.050 (0.022) | | | 0.020 |
| **Family Environment** | **Est. (SE)** | | | **P-value** | **Est. (SE)** | | | **P-value** |
| Family Conflict (FES) | 0.142 (0.033) | | | < 0.001 | 0.032 (0.024) | | | 0.178 |
| Perceived Stress (PSS) | 0.238 (0.049) | | | < 0.001 | 0.014 (0.031) | | | 0.651 |
| AL Positive Parenting | -0.063 (0.046) | | | 0.168 | 0.049 (0.028) | | | 0.082 |
| AL Parental Involvement | -0.194 (0.044) | | | < 0.001 | -0.020 (0.031) | | | 0.528 |
| AL Poor Monitoring | 0.166 (0.044) | | | < 0.001 | 0.006 (0.030) | | | 0.844 |
| AL Inconsistent Discipline | 0.217 (0.045) | | | < 0.001 | -0.014 (0.029) | | | 0.625 |
| **ADHD Polygenic Risk, Medication Status, and Sex** | **Est. (SE)** | | | **P-value** | **Est. (SE)** | | | **P-value** |
| ADHD PRS | 0.230 (0.032) | | | < 0.001 | 0.026 (0.026) | | | 0.301 |
| LT Med. Status | 0.642 (0.022) | | | < 0.001 | 0.174 (0.037) | | | < 0.001 |
| Sex | 0.162 (0.028) | | | < 0.001 | 0.006 (0.022) | | | 0.783 |
| **Cognition** | **Est. (SE)** | | | **P-value** | **Est. (SE)** | | | **P-value** |
| Cog. Speed (Composite) | 0.198 (0.03) | | | < 0.001 | 0.012 (0.026) | | | 0.639 |
| Working Memory (Composite) | -0.265 (0.029) | | | < 0.001 | -0.010 (0.024) | | | 0.639 |
| Cog. Arousal (Composite) | -0.276 (0.029) | | | < 0.001 | -0.010 (0.024) | | | 0.668 |
| SSRT | 0.197 (0.031) | | | < 0.001 | -0.012 (0.031) | | | 0.692 |
| **Outcomes** | **Odds Ratio** | **Lower CI** | **Upper CI** | **P-value** | **Odds Ratio** | **Lower CI** | **Upper CI** | **P-value** |
| Alcohol Use | 0.835 | 0.700 | 0.996 | 0.045 | 1.097 | 0.934 | 1.290 | 0.259 |
| Marijuana Use | 0.929 | 0.772 | 1.118 | 0.438 | 1.183 | 1.000 | 1.400 | 0.050 |
| Suicidality | 1.141 | 0.969 | 1.345 | 0.115 | 1.124 | 0.965 | 1.309 | 0.133 |

| | | | | | | | | |
|---|---|---|---|---|---|---|---|---|
| Mood Disorder Onset | 1.156 | 0.961 | 1.391 | 0.124 | 1.116 | 0.895 | 1.392 | 0.329 |
| Anxiety Disorder Onset | 1.184 | 0.978 | 1.435 | 0.084 | 0.875 | 0.683 | 1.123 | 0.294 |
| Disruptive Disorder Onset | 1.023 | 0.790 | 1.325 | 0.862 | 1.250 | 0.856 | 1.824 | 0.248 |
| Any New Disorder Onset | 1.141 | 0.963 | 1.352 | 0.129 | 1.016 | 0.832 | 1.242 | 0.873 |

Table S23. Associations between KSADS-measured inattention symptoms at baseline (left-most columns) and over time (right-most columns) in the Oregon-ADHD-1000 cohort, and predictor/outcome variables of interest.

##### *Oregon ADHD-1000 KSADS Hyperactivity.*

As with inattention, the results of the regression analyses conducted with KSADS-measured hyperactivity in the Oregon-ADHD-1000 cohort and our predictors and outcomes of interest agreed with those conducted with both Oregon-ADHD-1000 ADHDRS and ABCD KSADS hyperactivity measures. As expected, family/social environment and cognitive variables stood out as significant associations with baseline symptoms, but not with slope.

*Community Environment:*

*Baseline.* Following multiple testing corrections, one environmental predictor (ED) achieved statistical significance in relation to KSADS-measured hyperactivity at baseline (coeff: -0.09 (0.029), p = 0.002). A nominal effect was also observed for ADI (coeff = 0.074 (0.028), p = 0.008); see **Table S24.**

*Trajectory.* Following multiple testing corrections, no environmental predictors achieved statistical significance in relation to KSADS-measured hyperactivity over time.

*Family Environment:*

*Baseline.* All family and social predictors were significantly associated with KSADS - measured hyperactivity at baseline (p < 0.001; **Table S24**), except for positive parenting and poor monitoring.

*Trajectory.* No family and social predictors were significantly associated with KSADS - measured inattention over time.

*ADHD Polygenic Risk, Medication Status, and Sex:*

*Baseline.* Lifetime medication status (coeff = 0.524 (0.025), p < 0.001), polygenic risk (coeff = 0.230 (0.032), p < 0.001), and male sex (coeff = 0.183 (0.028), p < 0.001) were significantly associated with KSADS-measured hyperactivity at baseline; see **Table S24**.

*Trajectory.* Following multiple testing corrections, lifetime medication status, polygenic risk, and sex were not significantly associated with KSADS-measured hyperactivity over time.

*Cognition:*

Following multiple testing corrections, no cognitive predictors were significantly associated with KSADS -measured hyperactivity either at baseline or over time.

*Outcomes:*

Following multiple testing corrections, no outcomes were significantly associated with KSADS -measured hyperactivity either at baseline or over time.

| | **OR1000 KSADS Hyperactivity** | | | |
|---|---|---|---|---|
| | **Intercept** | | **Slope** | |
| **Community Environment** | **Est. (SE)** | **P-value** | **Est. (SE)** | **P-value** |
| ADI | 0.074 (0.028) | 0.008 | -0.006 (0.011) | 0.568 |
| ED | -0.090 (0.029) | 0.002 | 0.003 (0.009) | 0.736 |
| HE | 0.013 (0.03) | 0.656 | 0 (0.009) | 0.959 |
| SE | -0.034 (0.029) | 0.239 | 0.007 (0.009) | 0.453 |
| COI | -0.051 (0.029) | 0.079 | 0.007 (0.009) | 0.479 |
| NO2 | -0.008 (0.027) | 0.784 | 0.008 (0.007) | 0.301 |
| PM2.5 | 0.014 (0.028) | 0.606 | 0.012 (0.008) | 0.126 |
| Decile | -0.030 (0.028) | 0.293 | 0.012 (0.009) | 0.182 |
| **Family Environment** | **Est. (SE)** | **P-value** | **Est. (SE)** | **P-value** |
| Family Conflict (FES) | 0.119 (0.033) | < 0.001 | -0.008 (0.010) | 0.454 |
| Perceived Stress (PSS) | 0.222 (0.049) | < 0.001 | 0.006 (0.013) | 0.624 |
| AL Positive Parenting | 0.003 (0.046) | 0.949 | -0.006 (0.012) | 0.623 |
| AL Parental Involvement | -0.137 (0.044) | 0.002 | -0.016 (0.014) | 0.228 |

| | | | | | | | | |
|---|---|---|---|---|---|---|---|---|
| AL Poor Monitoring | 0.082 (0.046) | | | 0.074 | 0.005 (0.013) | | | 0.674 |
| AL Inconsistent Discipline | 0.192 (0.048) | | | < 0.001 | -0.004 (0.012) | | | 0.759 |
| **ADHD Polygenic Risk, Medication Status, and Sex** | **Est. (SE)** | | | **P-value** | **Est. (SE)** | | | **P-value** |
| ADHD PRS | 0.230 (0.032) | | | < 0.001 | 0 (0.010) | | | 0.980 |
| LT Med. Status | 0.524 (0.025) | | | < 0.001 | 0.027 (0.012) | | | 0.018 |
| Sex | 0.183 (0.028) | | | < 0.001 | 0.002 (0.009) | | | 0.817 |
| **Cognition** | **Est. (SE)** | | | **P-value** | **Est. (SE)** | | | **P-value** |
| Cog. Speed (Composite) | 0.105 (0.032) | | | 0.001 | 0.013 (0.011) | | | 0.255 |
| Working Memory (Composite) | -0.175 (0.032) | | | < 0.001 | -0.013 (0.010) | | | 0.229 |
| Cog. Arousal (Composite) | -0.199 (0.031) | | | < 0.001 | -0.013 (0.009) | | | 0.147 |
| SSRT | 0.166 (0.035) | | | < 0.001 | 0.005 (0.011) | | | 0.627 |
| **Outcomes** | **Odds Ratio** | **Lower CI** | **Upper CI** | **P-value** | **Odds Ratio** | **Lower CI** | **Upper CI** | **P-value** |
| Alcohol Use | 0.917 | 0.764 | 1.101 | 0.355 | 0.868 | 0.604 | 1.247 | 0.445 |
| Marijuana Use | 1.045 | 0.865 | 1.263 | 0.646 | 0.961 | 0.659 | 1.402 | 0.838 |
| Suicidality | 1.064 | 0.902 | 1.256 | 0.460 | 1.427 | 1.009 | 2.020 | 0.045 |
| Mood Disorder Onset | 1.055 | 0.878 | 1.267 | 0.570 | 1.304 | 0.793 | 2.144 | 0.296 |
| Anxiety Disorder Onset | 1.224 | 1.008 | 1.487 | 0.042 | 0.693 | 0.416 | 1.154 | 0.158 |
| Disruptive Disorder Onset | 0.860 | 0.653 | 1.133 | 0.284 | 1.569 | 0.666 | 3.694 | 0.303 |
| Any New Disorder Onset | 1.035 | 0.871 | 1.229 | 0.698 | 0.974 | 0.610 | 1.555 | 0.912 |

Table S24. Associations between KSADS-measured hyperactivity symptoms at baseline (left-most columns) and over time (right-most columns) in the Oregon-ADHD-1000 cohort, and predictor/outcome variables of interest.

#### ABCD

##### *ABCD CBCL ADHD.*

The results of the regression analyses conducted with CBCL-measured ADHD symptoms in the ABCD cohort and our predictors and outcomes of interest were in agreement with all other measures of inattention, within and between cohorts. Family/social environment and cognition stood out as significant effects with respect to baseline symptoms. Perceived stress was significantly associated with symptom slope, which differed from other measures, but with a consistent direction of effect. Mood and disruptive disorder onset were significantly associated with CBCL ADHD slope, also with consistent directions of effect.

*Community Environment:*

*Baseline.* No environmental predictors achieved statistical significance in relation to CBCL ADHD trait measures at baseline.

*Trajectory.* SE was significantly associated with CBCL ADHD trait measures over time (coeff = 0.06 (0.019), p = 0.001) and a nominal effect was observed for COI (0.054 (0.018), p = 0.003); see **Table S25.**

*Family Environment:*

*Baseline.* All family and social predictors were significantly associated with CBCL - measured ADHD traits at baseline (p < 0.001; **Table S25**) except for educational support (not significant), and neglectful behavior, which was nominally significant: coeff = -0.091 (0.031), p = 0.004; see **Table S25**).

*Trajectory.* Only parent PSS was significantly associated with CBCL-measured ADHD traits over time (coeff = 0.069 (0.018), p < 0.001; **Table S25**).

*ADHD Polygenic Risk, Medication Status, and Sex.*

*Baseline.* Lifetime medication status (coeff = 0.189 (0.031), p < 0.001) and polygenic risk (coeff = 0.142 (0.028), p < 0.001) were significantly associated with CBCL-measured ADHD traits at baseline, and male sex was nominally significant (coeff = 0.079 (0.028), p = 0.005); see **Table S25**.

*Trajectory.* Lifetime medication status, polygenic risk, and sex were not significantly associated with KSADS-measured hyperactivity over time.

*Cognition:*

*Baseline.* All cognitive predictors were significantly associated with CBCL -measured ADHD traits at baseline (p < 0.001; **Table S25).**

*Trajectory.* No cognitive predictors were significantly associated with CBCL -measured ADHD trait change over time.

*Outcomes:*

*Baseline.* The onset of new anxiety and disruptive disorders, and new disorder onset overall, were all significantly associated with CBCL-measured ADHD trait change at baseline ($p < 0.001$; **Table S25**). Mood disorder onset was nominally significant (OR = 1.822 [1.185 – 2.799], p = 0.007; **Table S25).**

*Trajectory.* The onset of new mood and disruptive disorders, and new disorder onset overall, were all significantly associated with CBCL-measured ADHD trait change over time ($p < 0.001$; **Table S25).**

| | **ABCD CBCL ADHD** | | | |
|---|---|---|---|---|
| | **Intercept** | | **Slope** | |
| **Community Environment** | **Est. (SE)** | **P-value** | **Est. (SE)** | **P-value** |
| ADI | 0.049 (0.028) | 0.080 | 0 (0.018) | 0.997 |
| ED | -0.028 (0.03) | 0.352 | 0.033 (0.019) | 0.081 |
| HE | -0.014 (0.029) | 0.626 | 0.028 (0.018) | 0.118 |
| SE | -0.014 (0.029) | 0.635 | 0.060 (0.019) | 0.001 |
| COI | 0.003 (0.029) | 0.909 | 0.054 (0.018) | 0.003 |
| NO2 | 0.009 (0.029) | 0.755 | 0.005 (0.019) | 0.788 |
| PM2.5 | 0.020 (0.028) | 0.477 | -0.003 (0.018) | 0.872 |
| Decile | -0.011 (0.029) | 0.703 | -0.033 (0.018) | 0.068 |
| **Family Environment** | | | | |
| Family Conflict (FES) | 0.319 (0.028) | < 0.001 | 0.015 (0.02) | 0.443 |
| Perceived Stress (PSS) | 0.212 (0.026) | < 0.001 | 0.069 (0.018) | < 0.001 |
| Positive Parenting (CRPBI Emotional Warmth) | -0.103 (0.029) | < 0.001 | -0.008 (0.018) | 0.672 |
| Involvement (MNBS Educational Support) | -0.041 (0.030) | 0.176 | -0.030 (0.019) | 0.122 |
| Neglectful Behavior (MNBS Overall Mean) | -0.091 (0.031) | 0.004 | -0.008 (0.019) | 0.698 |
| Parental Monitoring Questionnaire | -0.154 (0.029) | < 0.001 | -0.002 (0.019) | 0.928 |

| ADHD Polygenic Risk, Medication Status, and Sex | Est. (SE) | | | P-value | Est. (SE) | | | P-value |
|---|---|---|---|---|---|---|---|---|
| ADHD PRS | 0.142 (0.028) | | | < 0.001 | -0.001 (0.017) | | | 0.956 |
| LT Med. Status | 0.189 (0.031) | | | < 0.001 | -0.014 (0.018) | | | 0.439 |
| Sex | 0.079 (0.028) | | | 0.005 | 0.008 (0.018) | | | 0.650 |
| **Cognition** | **Est. (SE)** | | | **P-value** | **Est. (SE)** | | | **P-value** |
| Cog. Speed (NIH Toolbox Pattern Comparison Task) | -0.141 (0.029) | | | < 0.001 | -0.008 (0.019) | | | 0.656 |
| Working Memory (NIH Toolbox List Sorting Task) | -0.196 (0.027) | | | < 0.001 | 0.011 (0.018) | | | 0.543 |
| Response Inhibition (NIH Toolbox Flanker Task) | -0.121 (0.030) | | | < 0.001 | 0.019 (0.019) | | | 0.307 |
| SSRT | 0.139 (0.029) | | | < 0.001 | 0.011 (0.022) | | | 0.602 |
| **Outcomes** | **Odds Ratio** | **Lower CI** | **Upper CI** | **P-value** | **Odds Ratio** | Lower CI | Upper CI | **P-value** |
| Alcohol Use | 0.619 | 0.247 | 1.547 | 0.302 | 1.176 | 0.358 | 3.860 | 0.789 |
| Marijuana Use | 0.887 | 0.596 | 1.320 | 0.557 | 0.558 | 0.294 | 1.059 | 0.074 |
| Suicidality | 1.086 | 0.946 | 1.247 | 0.252 | 1.071 | 0.877 | 1.310 | 0.502 |
| Mood Disorder Onset | 1.822 | 1.185 | 2.800 | 0.007 | 3.421 | 2.045 | 5.725 | < 0.001 |
| Anxiety Disorder Onset | 1.777 | 1.295 | 2.438 | < 0.001 | 1.287 | 0.818 | 2.026 | 0.275 |
| Disruptive Disorder Onset | 2.213 | 1.778 | 2.754 | < 0.001 | 2.208 | 1.541 | 3.163 | < 0.001 |
| Any New Disorder Onset | 1.962 | 1.623 | 2.365 | < 0.001 | 1.973 | 1.466 | 2.655 | < 0.001 |

Table S25. Associations between CBCL-measured inattention symptoms at baseline (left-most columns) and over time (right-most columns) in the ABCD cohort, and predictor/outcome variables of interest.

*ABCD CBCL Inattention.*

The results of the regression analyses conducted with CBCL-measured inattention symptoms in the ABCD cohort and our predictors and outcomes of interest were in agreement with all other measures of inattention, within and between cohorts, and were virtually the same as those observed with respect to the CBCL ADHD measure.

*Community Environment:*

*Baseline.* No environmental predictors achieved statistical significance in relation to CBCL-measured inattention at baseline.

*Trajectory.* SE was significantly associated with CBCL-measured inattention over time (coeff = 0.055 (0.018), p = 0.002) and a nominal effect was observed for COI (0.051 (0.018), p = 0.005); see **Table S26**.

*Family Environment:*

*Baseline.* All family and social predictors were significantly associated with CBCL - measured inattention at baseline (p <= 0.001; **Table S26**), except for emotional development support.

*Trajectory.* No family and social predictors were significantly associated with CBCL - measured inattention over time, although a nominal effect was observed for parent PSS (coeff = 0.055 (0.019), p = 0.004; see **Table S26**).

*ADHD Polygenic Risk, Medication Status, and Sex.*

*Baseline.* Following multiple testing corrections, lifetime medication status (coeff = 0.193 (0.03), p < 0.001) and polygenic risk (coeff = 0.138 (0.028), p < 0.001) were significantly associated with CBCL-measured inattention at baseline; see **Table S26.**

*Trajectory.* Lifetime medication status, polygenic risk, and sex were not significantly associated with CBCL-measured inattention over time.

*Cognition:*

*Baseline.* All cognitive predictors were significantly associated with CBCL -measured inattention at baseline (p < 0.001; **Table S26).**

*Trajectory.* No cognitive predictors were significantly associated with CBCL -measured inattention over time.

*Outcomes:*

*Baseline.* The onset of new anxiety and disruptive disorders, and new disorder onset overall, were all significantly associated with CBCL-measured inattention at baseline (p <

0.001; **Table S26**). Mood disorder onset was nominally significant (OR = 1.580 [1.106 - 2.258], p = 0.008; see **Table S26**).

*Trajectory.* Following multiple testing corrections, the onset of new mood and disruptive disorders, and new disorder onset overall, were all significantly associated with CBCL-measured inattention over time (p < 0.001; **Table S26)**.

| | **ABCD CBCL Inattention** | | | |
|---|---|---|---|---|
| | **Intercept** | | **Slope** | |
| **Community Environment** | **Est. (SE)** | **P-value** | **Est. (SE)** | **P-value** |
| ADI | 0.047 (0.028) | 0.085 | -0.004 (0.017) | 0.830 |
| ED | -0.035 (0.03) | 0.242 | 0.033 (0.019) | 0.083 |
| HE | -0.017 (0.029) | 0.556 | 0.024 (0.018) | 0.178 |
| SE | 0.004 (0.029) | 0.882 | 0.055 (0.018) | 0.002 |
| COI | -0.005 (0.029) | 0.861 | 0.051 (0.018) | 0.005 |
| NO2 | 0.015 (0.029) | 0.607 | 0.001 (0.018) | 0.966 |
| PM2.5 | 0.029 (0.028) | 0.300 | -0.007 (0.018) | 0.711 |
| Decile | -0.005 (0.029) | 0.856 | -0.032 (0.018) | 0.067 |
| **Family Environment** | | | | |
| Family Conflict (FES) | 0.315 (0.028) | < 0.001 | 0.013 (0.019) | 0.478 |
| Perceived Stress (PSS) | 0.216 (0.026) | < 0.001 | 0.055 (0.019) | 0.004 |
| Positive Parenting (CRPBI Emotional Warmth) | -0.109 (0.029) | < 0.001 | -0.017 (0.019) | 0.374 |
| Involvement (MNBS Educational Support) | -0.051 (0.032) | 0.115 | -0.048 (0.019) | 0.010 |
| Neglectful Behavior (MNBS Overall Mean) | -0.113 (0.033) | 0.001 | -0.015 (0.019) | 0.449 |
| Parental Monitoring Questionnaire | -0.175 (0.029) | < 0.001 | -0.006 (0.019) | 0.742 |
| **ADHD Polygenic Risk, Medication Status, and Sex** | **Est. (SE)** | **P-value** | **Est. (SE)** | **P-value** |
| ADHD PRS | 0.138 (0.028) | < 0.001 | 0.002 (0.018) | 0.890 |
| LT Med. Status | 0.193 (0.030) | < 0.001 | -0.008 (0.018) | 0.667 |

| | | | | | | | | |
|---|---|---|---|---|---|---|---|---|
| Sex | 0.068 (0.029) | | | 0.018 | -0.004 (0.017) | | | 0.830 |
| **Cognition** | **Est. (SE)** | | | **P-value** | **Est. (SE)** | | | **P-value** |
| Cog. Speed (NIH Toolbox Pattern Comparison Task) | -0.168 (0.029) | | | < 0.001 | -0.012 (0.018) | | | 0.511 |
| Working Memory (NIH Toolbox List Sorting Task) | -0.203 (0.027) | | | < 0.001 | 0.008 (0.017) | | | 0.647 |
| Response Inhibition (NIH Toolbox Flanker Task) | -0.142 (0.030) | | | < 0.001 | 0.024 (0.017) | | | 0.161 |
| SSRT | 0.139 (0.028) | | | < 0.001 | 0.006 (0.021) | | | 0.762 |
| **Outcomes** | **Odds Ratio** | **Lower CI** | **Upper CI** | **P-value** | **Odds Ratio** | **Lower CI** | **Upper CI** | **P-value** |
| Alcohol Use | 0.665 | 0.282 | 1.568 | 0.354 | 1.277 | 0.378 | 4.313 | 0.694 |
| Marijuana Use | 0.880 | 0.581 | 1.332 | 0.541 | 0.538 | 0.283 | 1.025 | 0.060 |
| Suicidality | 1.056 | 0.922 | 1.208 | 0.427 | 1.102 | 0.900 | 1.350 | 0.348 |
| Mood Disorder Onset | 1.580 | 1.106 | 2.258 | 0.013 | 3.258 | 1.692 | 6.274 | < 0.001 |
| Anxiety Disorder Onset | 1.796 | 1.345 | 2.399 | < 0.001 | 1.418 | 0.907 | 2.217 | 0.125 |
| Disruptive Disorder Onset | 2.187 | 1.786 | 2.678 | < 0.001 | 2.301 | 1.639 | 3.231 | < 0.001 |
| Any New Disorder Onset | 1.934 | 1.625 | 2.300 | < 0.001 | 2.060 | 1.526 | 2.780 | < 0.001 |

Table S26. Associations between CBCL-measured ADHD symptoms at baseline (left-most columns) and over time (right-most columns) in the ABCD cohort, and predictor/outcome variables of interest.

##### *ABCD BPM Inattention (teacher-reported).*

The results of the regression analyses conducted with BPM-measured inattention symptoms in the ABCD cohort and our predictors and outcomes of interest broadly agreed with all other measures of inattention, within and between cohorts. Family/social environment and cognitive variables were, as expected, significantly associated with baseline symptoms but not change over time. However, the lifetime alcohol use outcome variable was significantly associated with BPM-measured inattention slope, a finding which did not emerge for any other measure, while the expected disorder onset outcomes (mood, disruptive disorders) were not significantly associated with BPM-measured inattention slope (**Table S27**).

*Community Environment:*

*Baseline.* Following multiple testing corrections, ED, HE, SE, and COI were all significantly associated environmental predictors with BPM-measured inattention at baseline ($p <= 0.002$; see **Table S27).**

*Trajectory.* No environmental predictors achieved statistical significance in relation to BPM-measured inattention over time.

*Family Environment:*

*Baseline.* Family conflict, emotional warmth, and parental monitoring were significantly associated with BPM-measured inattention at baseline, and a nominal effect was observed for neglectful behavior (**Table S27).**

*Trajectory.* No family or social predictors achieved statistical significance in relation to BPM-measured inattention over time.

*ADHD Polygenic Risk, Medication Status, and Sex.*

*Baseline.* Lifetime medication status (coeff = 0.164 (0.034), $p < 0.001$) and polygenic risk (coeff = 0.292 (0.03), $p < 0.001$) and male sex (coeff = 0.218 (0.029), $p < 0.001$) were significantly associated with BPM-measured inattention at baseline; see **Table S27.**

*Trajectory.* Lifetime medication status, polygenic risk, and sex were not significantly associated with BPM-measured inattention over time.

*Cognition:*

*Baseline.* All cognitive predictors were significantly associated with BPM -measured inattention at baseline ($p <= 0.001$; see **Table S27).**

*Trajectory.* No cognitive predictors were significantly associated with BPM -measured inattention over time.

*Outcomes:*

*Baseline.* New disruptive disorder onsets and new disorder onsets generally were significantly associated with BPM-measured inattention at baseline (OR = 1.693 [1.278-2.246], $p < 0.001$ and OR = 1.560 [1.264 – 1.926], $p < 0.001$ respectively) and a nominal effect was observed for anxiety disorder onset (OR = 1.560 [1.138 – 2.222], $p = 0.007$); see **Table S27.**

*Trajectory.* Alcohol use was significantly associated with BPM-measured shifts in attention over time (OR = 5.222, [5.1 – 5.347], $p = 0.002$; see **Table S27).**

| | ABCD BPM Inattention | | | |
|---|---|---|---|---|
| | Intercept | | Slope | |
| **Community Environment** | **Est. (SE)** | **P-value** | **Est. (SE)** | **P-value** |
| ADI | 0.080 (0.032) | 0.012 | 0.008 (0.017) | 0.658 |
| ED | -0.116 (0.033) | < 0.001 | -0.027 (0.016) | 0.089 |
| HE | -0.10 (0.033) | 0.002 | 0.003 (0.016) | 0.868 |
| SE | -0.102 (0.033) | 0.002 | -0.009 (0.015) | 0.539 |
| COI | -0.114 (0.033) | 0.001 | -0.013 (0.015) | 0.391 |
| NO2 | 0.004 (0.032) | 0.900 | -0.003 (0.015) | 0.836 |
| PM2.5 | 0.045 (0.032) | 0.161 | -0.007 (0.016) | 0.651 |
| Decile | 0.031 (0.032) | 0.334 | 0.024 (0.015) | 0.103 |
| **Family Environment** | | | | |
| Family Conflict (FES) | 0.215 (0.031) | < 0.001 | 0.011 (0.017) | 0.526 |
| Perceived Stress (PSS) | 0.042 (0.036) | 0.234 | -0.004 (0.016) | 0.807 |
| Positive Parenting (CRPBI Emotional Warmth) | -0.136 (0.034) | < 0.001 | -0.023 (0.015) | 0.130 |
| Involvement (MNBS Educational Support) | -0.028 (0.035) | 0.419 | -0.009 (0.013) | 0.479 |
| Neglectful Behavior (MNBS Overall Mean) | -0.102 (0.036) | 0.004 | -0.023 (0.015) | 0.128 |
| Parental Monitoring Questionnaire | -0.211 (0.033) | < 0.001 | 0.017 (0.017) | 0.324 |
| **ADHD Polygenic Risk, Medication Status, and Sex** | **Est. (SE)** | **P-value** | **Est. (SE)** | **P-value** |
| ADHD PRS | 0.292 (0.030) | < 0.001 | 0.030 (0.016) | 0.062 |
| LT Med. Status | 0.164 (0.034) | < 0.001 | -0.012 (0.017) | 0.489 |
| Sex | 0.218 (0.029) | < 0.001 | 0.004 (0.015) | 0.768 |
| **Cognition** | **Est. (SE)** | **P-value** | **Est. (SE)** | **P-value** |
| Cog. Speed (NIH Toolbox Pattern Comparison Task) | -0.173 (0.032) | < 0.001 | -0.033 (0.015) | 0.030 |

| | | | | | | | | |
|---|---|---|---|---|---|---|---|---|
| Working Memory (NIH Toolbox List Sorting Task) | -0.30 (0.028) | | | < 0.001 | -0.011 (0.017) | | | 0.523 |
| Response Inhibition (NIH Toolbox Flanker Task) | -0.170 (0.032) | | | < 0.001 | 0.001 (0.015) | | | 0.945 |
| SSRT | 0.127 (0.037) | | | 0.001 | 0.014 (0.016) | | | 0.378 |
| **Outcomes** | **Odds Ratio** | **Lower CI** | **Upper CI** | **P-value** | **Odds Ratio** | **Lower CI** | **Upper CI** | **P-value** |
| Alcohol Use | 0.932 | 0.474 | 1.836 | 0.839 | 5.222 | 1.791 | 15.231 | 0.002 |
| Marijuana Use | 0.961 | 0.639 | 1.446 | 0.848 | 1.084 | 0.529 | 2.219 | 0.825 |
| Suicidality | 1.047 | 0.898 | 1.221 | 0.560 | 0.992 | 0.713 | 1.381 | 0.961 |
| Mood Disorder Onset | 1.302 | 0.859 | 1.975 | 0.215 | 1.438 | 0.643 | 3.217 | 0.376 |
| Anxiety Disorder Onset | 1.590 | 1.138 | 2.222 | 0.007 | 0.821 | 0.515 | 1.309 | 0.408 |
| Disruptive Disorder Onset | 1.693 | 1.277 | 2.246 | < 0.001 | 1.028 | 0.569 | 1.858 | 0.928 |
| Any New Disorder Onset | 1.560 | 1.264 | 1.926 | < 0.001 | 0.937 | 0.631 | 1.393 | 0.749 |

Table S27. Associations between BPM-measured inattention symptoms at baseline (left-most columns) and over time (right-most columns) in the ABCD cohort, and predictor/outcome variables of interest.

#### References

Cordova, M. M., Antovich, D. M., Ryabinin, P., Neighbor, C., Mooney, M. A., Dieckmann, N. F., Miranda-Dominguez, O., et al. (2022). Attention-Deficit/Hyperactivity Disorder: Restricted Phenotypes Prevalence, Comorbidity, and Polygenic Risk Sensitivity in the ABCD Baseline Cohort. *Journal of the American Academy of Child and Adolescent Psychiatry*, *61*(10), 1273–1284. United States.

Ellis, L. K., & Rothbart, M. K. (2001). Revision of the early adolescent temperament questionnaire. *Poster presented at the 2001 biennial meeting of the society for research in child development, Minneapolis, Minnesota*.

Ho, D., Imai, K., King, G., & Stuart, E. A. (2011). MatchIt: Nonparametric Preprocessing for Parametric Causal Inference. *Journal of Statistical Software*, *42*(8), 1–28.

Kozlowski, M. B., Antovich, D., Karalunas, S. L., & Nigg, J. T. (2022). Temperament in middle childhood questionnaire: New data on factor structure and applicability in a child clinical sample. *Psychological assessment*, *34*(12), 1081–1092. United States.

Kozlowski, M. B., Morton, H. E., Nigg, J. T., & Karalunas, S. L. (2025). Reproducible structure with measurement invariance for the Parent-Report Early Adolescent Temperament Questionnaire: Findings from three independent samples. *Psychological assessment*, *37*(5), 214–226. United States.

Nigg, J. T., Gustafsson, H. C., Karalunas, S. L., Ryabinin, P., McWeeney, S. K., Faraone, S. V., Mooney, M. A., et al. (2018). Working Memory and Vigilance as Multivariate Endophenotypes Related to Common Genetic Risk for Attention-Deficit/Hyperactivity Disorder. *Journal of the American Academy of Child and Adolescent Psychiatry*, *57*(3), 175–182. United States.
